# PvGTSeq and PvCRiSP: two amplicon-based targeted sequencing panels for *Plasmodium vivax*

**DOI:** 10.1101/2025.11.04.25339464

**Authors:** Paulo C. Manrique-Valverde, Chloe M. Hasund, Katrina A. Kelley, Jorge-Eduardo Amaya-Romero, Myriam Arévalo-Herrera, Raphael Brosula, Jose Calzada, Stella M. Chenet, Vladimir Corredor, Angela M. Early, Gustavo Fontecha, David A. Forero-Peña, Sócrates Herrera, Justin T. Lana, Margaret Laws, Reza Niles-Robin, Nicanor Obaldia, Ana M. Santamaria, Philipp Schwabl, Sarah Auburn, Daniel E. Neafsey

## Abstract

*Plasmodium vivax* is the main cause of malaria outside of sub-Saharan Africa, and in many settings it presents significant challenges to malaria elimination efforts. Despite some control successes in the Americas, regional annual case counts of malaria have increased by over 25% between 2014 and 2023, largely driven by *P. vivax.* Genomic surveillance can play a key role in understanding the extent to which disease persistence represents indigenous transmission as opposed to introduction of new strains through migration, and whether specific variants evade control measures. Efforts to make *P. vivax* genomic surveillance more cost-effective have led to the development of targeted sequencing-based methods, which strike a varying balance between assay sensitivity and breadth/informativeness. We introduce two new highly sensitive multiplexed amplicon sequencing panels for *P. vivax*: PvGTSeq and PvCRiSP. PvGTSeq requires selective whole-genome amplification (sWGA) and contains 249 amplicons—36 for antimalarial resistance and 213 for population structure—optimized for Latin America but applicable to all continents. PvCRiSP features four highly polymorphic amplicons that operate without sWGA. Both panels use a single multiplex PCR with non-proprietary reagents, achieve ≥75% amplicon recovery at parasitemias as low as five parasites/μL, and PvCRiSP remains effective with low quality DNA. The evaluation of 137 technical replicates with the PvGTSeq panel showed high sequencing accuracy (error rate 3.85e-4% - 2.87e-3%), and both panels efficiently detected alleles from minority clones in simulated polyclonal infections. We validated both panels with samples from Colombia, Guyana, Honduras, Panama, and Venezuela, and performed in-silico assessments using data from 16 countries worldwide, confirming that these two panels have high power to discriminate samples and assign global geographic origin to imported cases. These panels will therefore be useful tools for *P. vivax* molecular surveillance in diverse geographic settings.

**Author Summary:** Genomic surveillance of malaria parasites has proven highly valuable for tracking the spatial and temporal distribution of drug resistance markers, measuring population connectivity, and differentiating locally transmitted vs. imported infections, among other use cases. We have designed and validated two Illumina-based multiplexed PCR amplicon sequencing panels for *P. vivax*: PvGTSeq and PvCRiSP. The PvGTSeq panel incorporates 249 amplicons, covering candidate genes associated with drug resistance as well as amplicons capturing polymorphic regions particularly useful for fine scale spatial resolution of parasite populations in Latin America. PvCRiSP consists of only four highly polymorphic and ultra-sensitive amplicons to estimate complexity of infection (COI) and identify instances of clonal transmission, without a costly and effort-intensive pre-amplification step. Both panels reliably generate genotypic data for patient samples with parasitemia levels as low as five parasites/μL and do not use proprietary reagents. These panels are suited for use in diverse geographic regions, with particular utility for studying *P. vivax* in the Americas—a region that is facing significant challenges in malaria control and elimination.

## Introduction

*Plasmodium vivax* is the main cause of malaria outside of Africa. Each year, 6-7 million cases are reported, and in the Americas this Plasmodium species causes 76% of all malaria cases.(1) Despite efforts to reduce malaria, *P. vivax* possesses biological characteristics that make it resilient to current control measures. Compared to *Plasmodium falciparum*, *P. vivax* has an earlier developmental commitment to transmissible sexual stages (2) and can persist as a dormant hypnozoite in the liver that can reactivate weeks or months after the primary infection.(3) In addition to these biological advantages, several factors complicate the control and elimination of this parasite in the Americas: the high frequency of submicroscopic and asymptomatic infections, (4) its prevalence in remote rural and border regions with limited health surveillance, (5) and inadequate patient compliance with radical cure treatment (7 to 14 days primaquine) to clear hypnozoites from the liver. (6)

Despite successful malaria control and elimination efforts in some countries across the Americas, there has been an increase in *P. vivax* cases since 2014—even in areas that previously reported fewer than 1,000 cases annually or where *P. falciparum* was the dominant parasite. (1) Although Venezuela has experienced the most dramatic increase in cases, (7) other countries such as Colombia, Costa Rica, Guyana, Ecuador, Guatemala, Honduras, Nicaragua, and Panama have also shown increases. (1) The drivers of *P. vivax* persistence and resurgence are complex, and control programs therefore require cost-effective tools to monitor these population changes and enable evidence-based decision-making.

Genomic surveillance can play an important role in this goal, and the development of targeted sequencing-based methods for *P. falciparum* has made this tool more cost- effective and easy to transfer to National Malaria Control Programs. (8–12) Recently, the use of genomic surveillance has expanded to *P. vivax* with a variety of use cases. (13–20) The most frequent use case has been characterizing population connectivity and distinguishing imported infections from those that are locally acquired. Other applications include characterizing candidate antimalarial resistance-related genes and potential blood-stage vaccine targets, as well as identifying amplicons to distinguish between infection relapses, recrudescences, and reinfections.

Here, we introduce two new amplicon panels for targeted Illumina sequencing of *P. vivax*. The first, PvGTSeq, is a multipurpose panel composed of 249 amplicons, including 213 highly heterozygous amplicons for assessing parasite diversity, relatedness, and population structure at three geographic scales: global, intra-continental, and sub-national in Latin America and the Caribbean. This panel has enhanced sensitivity for identifying imported infections in the Americas, while maintaining utility for malaria molecular surveillance worldwide. Additionally, this panel includes 36 amplicons targeting ten candidate genes associated with antimalarial resistance. The second panel, PvCRiSP, consists of four highly-polymorphic amplicons. This high-sensitivity panel is suited towards efficient estimation of the number of genetically distinct parasites within a single infection (complexity of infection, or COI) and for identifying instances of clonal transmission, for example in localized outbreaks. Both panels work on all Illumina sequencing platforms and require a single multiplex PCR reaction prior to the indexing reaction. Both also involve non-proprietary and easily-sourced reagents, facilitating global access and sustainable programmatic use. In this manuscript, we describe the performance of the panels to assess their utility for various use cases.

## Methods

### Clinical samples and genomic databases

Multiple genomic data sources and clinical samples were used for the selection, standardization, and validation of PvGTSeq and PvCRiSP amplicons. In the selection process for PvGTSeq amplicons, genomic data from three different sources were used:

1) 1895 genomes from the MalariaGen Pv4 database, (21) 2) 705 genomes obtained from Guyana not present in MalariaGen Pv4, (22) and 3) 532 new genomes obtained from clinical samples from Colombia (200), Peru (90) and Venezuela (342). For the validation of PvGTSeq and PvCRiSP, a total of 821 additional clinical blood spot samples from Colombia (259), Guyana (460), Honduras (39), Panama (53), and Venezuela (10) were genotyped using one of these methods (Fig S1 in S1 Text). All sequenced clinical samples were collected with ethical approval from local institutions and their analysis was approved by the Harvard University Longwood Institutional Review Board (Protocols: IRB19-1779, IRB20-1636, IRB23-0621, IRB23-1567).

### Parasite DNA quantification

We measured parasitemia for a subset of clinical samples used in assay optimization, limit of detection measurement, and minor clone sensitivity analysis. Parasitemia was measured by real-time quantitative polymerase chain reaction (qPCR) using the 18S ribosomal gene (23) and a standard curve from the 3D7 strain of *P. falciparum*. Details about the master mix and cycling conditions are in S2 Text. Due to the lack of a quantified *P. vivax* culture or large volumes of a high-parasitemia sample that could be used as a control for all assays in this study, to quantify *P. vivax* parasitemia on clinical samples we used genomic DNA from the monoclonal *P. falciparum* reference strain, 3D7, as a quantitation standard. We prepared a positive control template representing infected human blood with 10,000 parasites/μL by mixing human genomic DNA (13.76 ng/μL) with 3D7 genomic DNA (0.92 ng/μL) at 2.66:1 ratio (v/v), yielding 10 ng/μL human gDNA and 0.25 ng/μL 3D7 gDNA. The 0.25 ng represents 10,000 *P. falciparum* genomes (based on 23-Mbp genome size and 660 g/mol bp average mass), assuming one haploid genome per infected cell. We created controls representing 10,000, 1000, 100, 10 and 1 parasites/μL by serial 1:10 dilution with 10 ng/μL human gDNA. Because primer pair amplification efficiency may differ between *P. falciparum* and *P. vivax*, we measured the amplification efficiency in the logarithmic PCR phase for each sample as a correction factor relative to the standard curve efficiency.

### Selection of amplicons and primer design

To describe patterns of population structure and connectivity at three geographic scales (global, Latin America, and subnational within Latin America), we selected discriminatory amplicons using Discriminant Analysis of Principal Components (DAPC). (24) Whole genome sequencing information (WGS) from the MalariaGEN Pv4 dataset (25) and lab-generated WGS were filtered to remove low-quality regions and genotyping error-prone loci. Genomes were divided into 150 bp segments, with segment selection based on: 1) >0.1% contribution to top discriminant components, 2) highest contribution within 200,000 bp windows, and 3) location in functionally annotated genes. Segments in uncharacterized genes were selected only when no other segments met the first criterion within a window. We performed this selection process independently for each of the three geographical scales, generating three subsets of segments. Segments shared across all three subsets were prioritized for primer design. For surveillance of antimalarial drug resistance, we identified 150 bp segments (with 50 bp flanking regions) covering polymorphic sites in 11 candidate resistance genes: *CRT* (PVP01_0109300), (26–32) *MRP1* (PVP01_0203000), (33–36) *DMT2* (PVP01_0312700), (35) *DHFR* (PVP01_0526600), (30,33,35,37–42) *MDR1* (PVP01_1010900), (27,33,42–47) *PI3K* (PVP01_1018600), (46,48) *ABCE1* (PVP01_1103800), (32) *KELCH13* (PVP01_1211100), (30,35,49) *MDR2* (PVP01_1259100), (50,51) *DHPS* (PVP01_1429500), (28,30,41,42,51–55) and *MRP2* (PVP01_1447300). (35,50) Finally, we contracted the services of GTseek LLC (56) to design multiplexable primer sets following GT-seq protocol specifications. (50) PvCRiSP consists of a subset of just four amplicons selected from the larger PvGTSeq panel.

These four amplicons were chosen based on their heterozygosity, ability to detect polyclonal infections, and superior amplification efficiency across the various Latin American countries analyzed with PvGTSeq.

### Whole genome sequencing and variant call

For clinical samples from Colombia, Guyana, and Venezuela we extracted total genomic DNA from dried blood spot samples (20–35 mm^2^ spotted area punched per sample) using KingFisher Ready DNA Ultra 2.0 Prefilled Plates on the Kingfisher Flex instrument (ThermoFisher Scientific). For Honduras, Panama, and Peru, parasite DNA was extracted using various other methods. We subsequently applied selective whole genome amplification (sWGA) to DNA extracts, (58) each 50 μL sample reaction consisting of 5 μL 10x phi29 polymerase buffer (NEB B0269S), 0.125 μL (2.5 μg) recombinant albumin (NEB B9200S), 0.5 μL primer mix (10 oligos combined at 250 μM, see below), 5 μL 10 mM dNTPs (Thermo Scientific), 16.375 nuclease-free water, 3 μL (30 units) phi29 DNA polymerase (M0269L), and 20 μL DNA. Reactions were prepared on ice, with components added in the order shown. We used ‘Pvset1’ for *P. vivax* as reported by Cowell et. al. 2017. Amplifications were generated using step down incubation (35 °C for 5 min, 34 °C for 10 min, 33 °C for 15 min, 32 °C for 20 min, 31 °C for 30 min, 30 °C for 16 h, and 65 °C for 15 min), followed by cooling to 4 °C. We subsequently slightly modified this protocol to enhance its sensitivity for low parasitemia samples (S2 Text). We then applied AMPure XP magnetic beads (Beckman Coulter A63881) at room temperature to exchange post-reaction sample buffer to 10 mM Tris- HCl + 0.1 mM EDTA. Final library construction using the NEBNext Ultra II FS DNA Library Prep Kit (NEB E6177) and 2 × 151 bp sequencing on the Illumina NovaSeq 6000 platform was completed at the Broad Institute. We aligned reads to the *P. vivax* P01 reference genome assemblies using BWA-MEM v0.7.17-r1188 (59) and called SNPs and INDELs using GATK v3.5-0-g36282e4 (60) ‘HaplotypeCaller’ and ‘GenotypeGVCFs’ according to best practices defined by the Pf3k consortium (http://www.malariagen.net/data_package/pf3k-5/). The mapping and joint variant call process also included samples from the MalariaGEN Pv4 dataset. All downstream genetic analyses focused on variant sites (SNPs and INDELs) in core regions of the genome. Samples missing genotype calls for >25% variant sites at ≥2% minor allele frequency (after application of the core region filter) were also excluded from further analysis. Following sample exclusions, we removed variant sites located in coding regions prone to alignment errors (coding regions with observed heterozygosity above the 95th percentile and variant site density exceeding the 92.5th percentile of all coding regions in all populations). We also removed variant sites located in homopolymers and di-nucleotide short tandem repeats, as these are prone to PCR errors.

### Panel protocols and optimization

To facilitate standardization of library generation procedures, we started with the conditions described for a previously developed *P. falciparum* protocol. (11) From there, we made modifications to annealing temperature and primer concentrations, the final volume of the multiplex PCR reaction, and the primer sets and enzymes for sWGA. These modifications were applied to improve sequencing yield in terms of the number of on-target reads per sample per amplicon. For this standardization, we selected 60 clinical samples from Colombia (30), Peru (15), and Venezuela (15), each with parasitemia levels above 100 parasites/μL. The final protocol for the library generation and sequencing for PvGTSeq and PvCRiSP can be found in S2 Text and S3 Text, respectively. Individual protocol modifications were evaluated in a paired manner, using the previous protocol conditions as a reference. Comparisons were made in terms of the number of reads per sample per amplicon, the total number of reads per sample, and the percentage of amplicons amplified per sample. Additionally, at each step, the rate of non-target product generation for each primer was evaluated, and the primer pairs that generated a higher amount of non-target reads compared to specific reads were removed from the protocol for subsequent optimization rounds (Fig S1 in S1 Text).

### Amplicon data analysis

We used the pipeline previously described for *P. falciparum* with some modifications to generalize its use for *P. vivax*. (11) Briefly, we processed paired-end Illumina sequencing data in the form of FASTQ files using a custom analysis pipeline for which documentation can be found at https://github.com/broadinstitute/malaria-amplicon-pipeline. This pipeline utilizes the Divisive Amplicon Denoising Algorithm (DADA2) (61) to obtain microhaplotypes (i.e., alleles or ‘amplicon sequence variants’). We first aligned microhaplotypes obtained from dada2 against a custom-built database of PvP01 reference sequences for each amplicon locus. We then summarized observed sequence polymorphism into a concise format by converting individual microhaplotypes into ‘pseudo-CIGAR’ strings using a custom python script that can be found at https://github.com/Paulonvnv/MHap-Analysis, The rules used for this conversion are also detailed in Fig S2 in S1 Text. Next, microhaplotypes were discarded if supported by fewer than 10 read-pairs or by less than 1% of total read-pairs within a heterozygous locus. We also filtered out microhaplotypes exhibiting less than 80% identity to the reference sequence. We then masked variants in both the FASTA sequence of the microhaplotype and its pseudo-cigar string representation according to three criteria: 1) SNPs or INDELs located in homopolymer regions (≥5 consecutive identical nucleotides), 2) INDELs positioned at either the beginning or end of the microhaplotype sequence, and 3) variants predominantly (> 66%) present as minor alleles and mainly found in heterozygous sites (> 66%) across the population. All downstream analyses were performed based on the pseudo-cigar strings using custom R scripts that can be found at https://github.com/Paulonvnv/PvGTSeq_PvCRiSP_paper. All functionalities created to handle pseudo-cigar strings are documented at https://github.com/Paulonvnv/MHap-Analysis. This pipeline is also implemented in a Terra workspace (see S1 Text)—a user-friendly cloud-based platform for processing amplicon sequencing data.

### Genotyping performance: amplification rate and limit of detection

Following pseudo-cigar generation and filtration steps, we evaluated the amplification rate and limit of detection (LOD) using a set of 160 and 240 samples for PvGTSeq and PvCRiSP, respectively, and from three different endemic countries: Colombia, Guyana, and Venezuela. For PvGTSeq, parasite concentrations ranged from 0.26 to 1309.38 parasites/μL, while for PvCRiSP parasite concentrations ranged from 0.03 to 130.94 parasites/μL respectively. The amplification rate was analyzed at both the amplicon and sample level, and was defined as the percentage of samples with ≥10 reads per amplicon and as the percentage of amplified amplicons per sample, respectively. The LOD was calculated for each individual amplicon as well as the complete panel of amplicons that pass the optimization step (249 for PvGTSeq and 4 for PvCRiSP). For each amplicon, the LOD was defined as the minimum parasite concentration needed to generate a microhaplotype with ≥10 reads with a 75% probability in the case of PvGTSeq, or 95% in the case of PvCRiSP. LOD at the level of the amplicon was calculated through logistic regression with a quasi-binomial distribution, where parasite concentration was the explanatory variable and success (1) or failure (0) of amplification in the samples was the response variable. The quasi-binomial distribution helped adjust for outliers caused by poor DNA quality or other factors that increase data dispersion not included in the model. Amplicons were classified as highly sensitive if they amplified samples with concentrations <10 parasites/μL, and only this set of amplicons was used for subsequent analysis.

The LOD at panel level was defined as the minimum parasite concentration needed to detect ≥10 reads per microhaplotype at 75% of all amplicons in the case of PvGTSeq and 100% of all amplicons in the case of PvCRiSP. In both cases, LOD was calculated through linear regression between the logarithm of parasite DNA concentration (explanatory variable) and the logarithm of the odds of amplified amplicons per sample (dependent variable).

### Consistency across assays

The consistency of genotyping was determined through reproducibility of the microhaplotype calls between technical replicates of monoclonal infections within the same sequencing run (30 duplicate samples), between sequencing runs (68 duplicate samples), and between genotypes obtained from amplicon sequencing and genotypes extracted from WGS VCF files (39 duplicate samples). Due to differences in filtering parameters for sequencing and alignment errors between GATK and our dada2-based pipeline, no quality filters were applied to the VCF files obtained from joint calling.

Instead, VCF files were reconverted into FASTA sequences that were incorporated into our analysis pipeline as if they were reads obtained from amplicon sequencing. Thus, for each technical replicate we calculated the error rate of the sequencing, defined as the percentage of nucleotide differences between replicates with respect to the total number of base pairs across all amplicons.

### Detection of major and minor clones in polyclonal infections

The ability of PvGTSeq and PvCRiSP to detect microhaplotypes from different clones that co-occur within the same infection at different concentrations was measured using mock polyclonal samples generated by combining samples from different geographical areas. Four samples were selected, two from Colombia and two from Guyana, with similar parasite concentrations (11.33 - 58.46 parasites/μL). Mixtures were made at seven different proportions (1:1, 1:2, 1:5, 1:10, 10:1, 5:1, and 2:1), with two biological replicates and two technical replicates. In all mock samples, the parasite concentration of the minority clone was always above the detection limit of the technique, so that the absence of detection should only be explained by competition between alleles during the amplification or sequencing steps.

### Diversity and utility of the amplicon panels

We subsequently evaluated the ability of both amplicon panels to discriminate whether two infections are identical by state or not in different geographic regions. This analysis only included populations, world regions, or countries with 20 or more monoclonal samples with coverage for at least 75% of the amplicons in the PvGTSeq panel. The genetic sequences of monoclonal samples were obtained from three different sources: 1) WGS from the MalariaGEN Pv4 database, 2) WGS from our laboratory, and 3) amplicon sequencing data from samples sequenced using the PvGTSeq amplicon panel. A monoclonal sample was defined as one that had an *F_ws_* score greater than or equal to 0.975 (62) and a fraction of heterozygous loci less than or equal to 0.05, with loci being defined as variant sites in the case of a VCF file for WGS samples, or as microhaplotypes in the case of samples sequenced by PvGTSeq. To maintain consistency in variant calling between WGS and PvGTSeq sequences, VCF files were converted into FASTA sequences and incorporated into our analysis pipeline as if they were reads obtained from amplicon sequencing.

To determine the diversity of the amplicons in each geographic region, each amplicon was described in terms of the 1) number of polymorphic sites, 2) the number of microhaplotypes, and 3) the per-site nucleotide diversity. The nucleotide diversity, or π, was defined as the average number of nucleotide differences per site between two randomly selected sequences from the population. All these metrics were described independently for the set of amplicons used for geographic attribution at different administrative levels, for amplicons of PvCRiSP and for amplicons covering candidate drug resistance loci.

We measured the capacity of PvGTSeq and PvCRiSP to distinguish between 1) parasites with distinct genomic sequences and 2) parasites belonging to distinct clonal groups. To assess discrimination of genomically distinct parasites, we used WGS data from genetically distinct monoclonal samples (we kept samples with pairwise IBS < 0.999 or > ∼10 SNPs difference) and samples sequenced only by PvGTSeq. For samples sequenced exclusively by PvGTSeq, we assumed all haplotypes were genomically distinct since they originated from different infected patients. For assessing whether parasites belong to distinct clonal groups, we relied solely on WGS information, defining a clonal group as haplotypes with genetic similarity (IBS) ≥ 0.99 or < 100 SNPs difference in pairwise comparisons. For both scenarios and both amplicon panels, the discriminatory power was quantified as the proportion of in silico pairwise sample comparisons from the same population yielding identity by state (IBS) values less than one. IBS was calculated as one minus the Hamming distance between sample pairs across all amplicons in each panel.

### Population Structure and Geographic attribution

We evaluated the ability of PvGTSeq to detect population structure at three geographic levels (global, intra-continental, and sub-national). At the global and intra-continental levels, population structure was assessed through principal coordinate analysis (PCoA), while at the sub-national level, a network analysis was used instead. For both approaches, pairwise genetic similarity between infections was determined using IBS. To avoid introduction of bias due to natural selection,(63) amplicons from candidate drug resistance loci were excluded from this analysis.

### Data availability

The sequence data generated by this study has been deposited in the NCBI Sequence Read Archive (https://www.ncbi.nlm.nih.gov/sra) under BioProject PRJNA1092690.

Data preparation scripts are available at https://github.com/Paulonvnv/PvGTSeq_PvCRiSP_paper. All other data are included in the manuscript and supporting information.

## Results

### Selection of amplicons and protocol optimization

We identified 60,056 variant sites meeting our inclusion criteria: located in coding regions not prone to alignment errors, not in homopolymers or di-nucleotide short tandem repeats. A total of 1,292 genomes had coverage ≥ 75% supported by ≥ 5 reads for these variant sites (Fig S1 in S1 Text). Using DAPC, we identified 263 segments (150 bp each) contributing to population differentiation across three geographic scales (Fig S1 in S1 Text). We also identified 57 non-overlapping segments containing polymorphisms in 11 genes previously associated with antimalarial resistance. From this pool of 320 segments, we successfully designed primers for 271, which underwent successive rounds of protocol optimization. Of these, 249 amplicons amplified efficiently and specifically in the 60 clinical samples used for the standardization step. The final protocol for library generation and sequencing is detailed in the S2 Text and S3 Text, and the complete list of 249 amplicons, including primer information and reference genome coordinates, is provided in the S4 Table. The finalized panel consists of 213 amplicons specialized for geographic differentiation (112 for global-scale, 122 for between-country, and 110 for subnational differentiation in Latin America). Note that many of the amplicons nevertheless function at multiple geographic scales. The finalized panel additionally targets 36 amplicons within 10 genes associated with antimalarial resistance. The segment identified for the *CRT* gene failed at the primer design stage. The number of amplicons for each candidate antimalarial resistance gene, and the coding regions covered by those amplicons are shown in Table 1; and the haplotype frequencies of these 10 genes are shown in S5 Table and in Fig S3 in S1 Text. Additionally, we selected four highly polymorphic amplicons from this panel that were heterozygous in most polyclonal samples and showed high amplification efficiency. These four amplicons—**C**G2_related, **RI**PR, VP**S**11, and **P**IGM—form the PvCRiSP panel. All amplicons are named according to their corresponding genes.

**Table 1:** Nucleotides and amino acids in antimalarial resistance candidate genes covered by PvGTSeq.

| Gene ID | Gene name | Number of amplicons | Covered nucleotides in the coding sequence | Covered amino acids |
| --- | --- | --- | --- | --- |
| PVP01_0203000 | *MRP1* | 7 | 339-431, 547-609, 1329-1509, 1980-2150, 2320-2478, 3199-3252, 4320-4427 | 113-144, 183-203, 443-503, 660-717, 774-826, 1067-1084, 1440-1476 |
| PVP01_0312700 | *DMT2* | 1 | 655-663 | 219-221 |
| PVP01_0526600 | *DHFR* | 1 | 17-72 | 6-24 |
| PVP01_1010900 | *MDR1* | 5 | 1147-1193, 1305-1479, 1646-1698, 2789-2903, 3677-3754 | 383-398, 435-493, 549-566, 930-968, 1226-1252 |
| PVP01_1018600 | *PI3K* | 10 | 560-610, 856-956, 1727-1770, 1898-1955, 2509-2656, 2780-2909, 3980-4035, 4262-4426, 4703-4754, 4951-5074 | 187-204, 286-319, 576-590, 633-652, 837-886, 927-970, 1327-1345, 1421-1476, 1568-1585, 1651-1692 |
| PVP01_1103800 | *ABC-E1* | 2 | 50-106, 1420-1475 | 17-36, 474-492 |
| PVP01_1211100 | *K13* | 1 | 1838-1900 | 613-634 |
| PVP01_1259100 | *MDR2* | 4 | 907-994, 1394-1565, 2525-2578, 3226-3288 | 303-332, 465-522, 842-860, 1076-1096 |
| PVP01_1429500 | *DHPS* | 3 | 207-290, 983-1040,<br>2028-2071 | 69-97, 328-347,<br>676-691 |
| PVP01_1447300 | *MRP2* | 2 | 5478-5592, 5745-5849 | 1826-1864,<br>1915-1950 |

### Genotyping performance

The amplification rate of the amplicons and the limit of detection varied among the three populations analyzed with PvGTSeq due to sample quality, but PvCRiSP yielded consistently high performance across populations. Regarding PvGTSeq, in Colombia, 190 out of the 249 amplicons amplified in more than 75% of the samples (Fig S4 A-B in S1 Text and S6 Table), with a median read depth of 95.5 (interquantile range or IQR = 409) across the 249 amplicons. The median LOD of these amplicons analyzed individually was 4.64 (IQR = 14.6) (Fig S4 C-D in S1 Text), and the minimum parasite concentration (LOD) to amplify 75% of the total 249 amplicons was 4.05 parasites/μL (95% CI from 2.5 to 5.9) (Fig 1). When considering only the amplicons that worked in more than 75% of the samples in Colombia, the LOD to amplify 95% of these 190 amplicons was 3.24 parasites/μL (95% CI from 1.18 to 6.15) (S7 Table). A similar result was obtained with samples from Guyana, where 201 amplicons amplified in more than 75% of the processed samples; however, parasite concentrations were above 5.73 parasites/μL, and LOD could not be determined either at the individual amplicon level or for the complete panel. Furthermore, there were 201 amplicons that had amplification rates above 75% in Colombia or Guyana. In contrast, Venezuelan samples showed poor amplification success, where only 21 amplicons were amplified in more than 75% of the samples.

**Fig 1:**
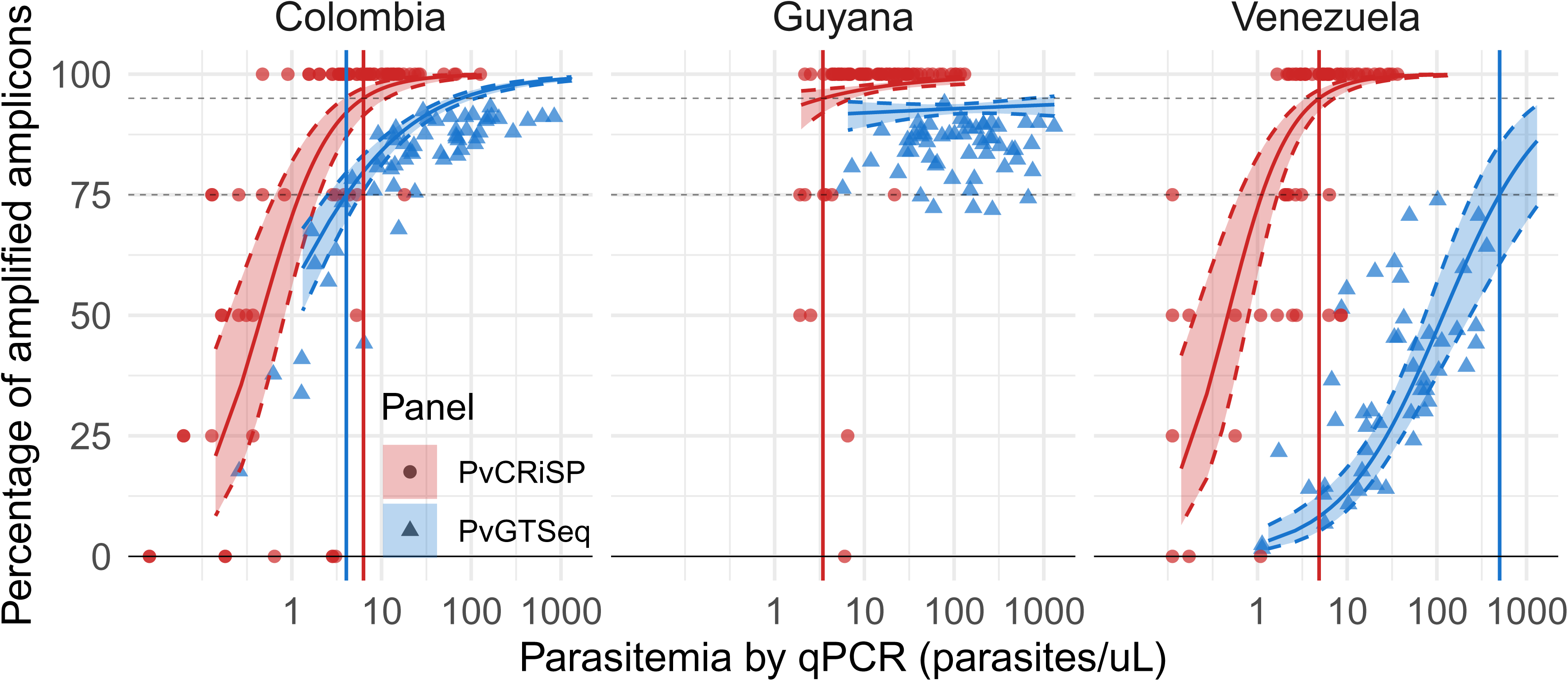
Limit of detection of PvGTSeq and PvCRiSP in Colombia, Guyana and Venezuela. The percentage of amplified amplicons per sample as a function of parasite concentration is illustrated for PvGTSeq (Blue) and PvCRiSP (Red). Each dot represents an individual sample and regression model (solid curve line) was constructed using the logarithm of parasite concentration (independent variable) and the logarithm of the odds of the amplified amplicons per sample (dependent variable) in each country. Vertical lines represent the minimum concentration a sample must have to amplify 75% of the 249 amplicons in PvGTSeq (blue lines) or the four amplicons of PvCRiSP (red lines). The shaded area between the dashed curve lines represents the 95% confidence interval.

We next investigated reproducibility, ability to detect minor alleles in polyclonal infections and population structure signals using the group of 201 amplicons that showed over 75% amplification success in Colombia or Guyana to avoid bias from missing data.

The PvCRiSP mini multiplex exhibited similar amplification success and LOD among the three populations. The four amplicons amplified on average more than 86% of the samples and had a median read depth of 131 (IQR = 446). The amplicon *CG*2_related_ showed the lowest amplification rate (77%) and the lowest read depth (median = 45.5, IQR = 127.25) across the three populations (S8 Table). The minimum concentration needed to amplify the four amplicons was 6.33 parasites/μL (95% CI from 4.92 to 8.23) and no significant differences were observed between the three countries when comparing their confidence intervals (S7 Table).

### Consistency across sample replicates

The results showed high reproducibility between technical replicates of monoclonal infections. Within the same sequencing run, of the 30 technical replicates analyzed, only in three cases were there differences between any of the amplicons between replicates. In these three cases, the difference was only one nucleotide between replicates. In two cases, the discrepancies were found in a marker against the gene *RAMA* (PVP01_0107500). In both of these two cases one of the replicates had a haplotype similar to the reference strain PvP01 while the other replicate contained only one non- synonymous polymorphism with respect to the reference. These two polymorphisms in the coding sequence were c.1797C>A (coding sequencing position 1797; amino acid change N599K) and c.1801G>A (G601R), and they were present in 58 and 43 samples respectively in our group of samples used for the estimation of LOD, an indication of cross-contamination. The third discrepancy was the nonsynonymous polymorphism c.536C>A (A179E) in the gene *SRPR alpha*, (PVP01_1105400) and this polymorphism was a singleton in the data set. The error rate including only the genotyping error was 3.85e-4% (phred Q34; permutation test, 95% CI 7.93e-5% to 1.12e-3%).

When comparing replicates between different sequencing runs, we observed that 16 out of the 68 replicates had a different microhaplotype. Most discrepant cases consisted of a single nucleotide polymorphism (11 cases). Other discrepant cases included the occurrence of two SNVs in a single amplicon (1 case), one INDEL plus one SNV in a single amplicon (2 cases), one INDEL and one SNV in two different amplicons (1 case), and one INDEL plus three SNVs in a single amplicon (1 case). Moreover, in five cases the discrepancy was in an amplicon that was heterozygous in one of the two replicates. Based on these observed discrepancies between replicates that were not likely caused by contamination the error rate was 1.42e-3% (phred Q28.5; 95% CI from 8.53e-4% to 2.21e-3%).

PvGTSeq panel replicates showed greater differences when compared with WGS replicates (Fig S53 in S1 Text). These discrepancies were explained by amplicons targeting seven genes containing INDELs whose representation was inconsistent in the variant calling process between WGS and amplicon sequencing. Excluding these markers, the error rate was 2.87e-3% (95% CI from 1.75e-3% to 4.44e-3%).

### Detection of major and minor clones in polyclonal infections

Next, we tested the utility of the 201 highly sensitive amplicons from PvGTSeq and the four highly sensitive amplicons of PvCRiSP for detecting minor alleles in mock polyclonal infections. The selected clinical samples for generating mock samples were the following: SP0112286092, G4GWM400, SP0112286092, and G4GWM400; henceforth we will refer to them as samples S1, S2, S3, and S4 respectively. Samples S1 and S3 were from Colombia and their concentrations were 198.25 and 203.05 parasites/μL, respectively. Samples S2 and S4 were from Guyana and their concentrations were 114.78 and 119.06 parasites/μL, respectively. When analyzing these samples individually through the panel of 201 amplicons from PvGTSeq, samples S1, S2, and S3 were monoclonal, while sample S4 was polyclonal, with a fraction of heterozygous amplicons equal to 0.22 and a maximum number of microhaplotypes per amplicon of 3. The Hamming distance between the haplotypes of samples S1 and S2 was 0.52, while for samples S3 and S4 it was 0.48. The mock sample mixtures generated with samples S1 and S2 were named Mock Sample 1 (MS1), and those generated from samples S3 and S4 were named MS2. Each combination was performed at seven different proportions between the samples (1:10, 1:5, 1:2, 1:1, 2:1, 5:1, and 10:1) and in duplicate, generating 28 mock samples in total. Additionally, the combinations were performed at a total parasitemia of 100.33 parasites/μL for MS1 and 58.46 parasites/μL for MS2 such that the concentration of the minority clone was always above 5.85 parasites/μL, therefore the absence of amplification of the clone is expected to reflect competition between alleles in PCR and/or sequencing, not the clone occurring below panel detection limits.

The PvGTSeq and PvCRiSP panels identified all mock samples as polyclonal, including those samples in which the major clone was in a 10:1 ratio compared to the minor clone (Fig 2). In PvGTSeq, the percentage of private alleles detected was >76% whenever the major clone was in a proportion of 1:1, 2:1, 5:1, or 10:1 relative to the other clone present in the sample. However, for the minor clone, the detection rate of its private alleles decreased linearly with respect to its proportion in the sample (Fig 2A). Thus, only when both clones were in similar proportions in the sample was the observed fraction of heterozygous loci close to the expectation (75%) (Fig S6A in S1 Text), and as the ratio between the major clone to the minor clone increased, the ratio between the observed and expected fraction of heterozygous loci decreased, reaching only 30% when the major:minor clone was in a 10:1 ratio. Additionally, the read depth of the major clone was consistently higher than the read depth of the minor clone across all analyzed amplicons and all technical and biological replicates (Fig S7 in S1 Text). Thus, a positive correlation was observed between the median ratio of read depth of private alleles between major and minor clones with respect to the ratio of DNA input concentrations per clone (Fig S6C in S1 Text). In the case of PvCRiSP, the detection rate of private alleles was always above 67%, even when the major:minor clone ratio was 10:1 (Fig 2B). Therefore, the ratio between observed and expected fraction of heterozygous loci was always one regardless of the proportion of clones within the mock sample (Fig S6B in S1 Text). This result is similar to that obtained with the 4CAST panel designed for *P. falciparum*, where the sensitivity to detect alleles from the minor clone was 94% even with ratios of 10:1. (11)

**Fig 2:**
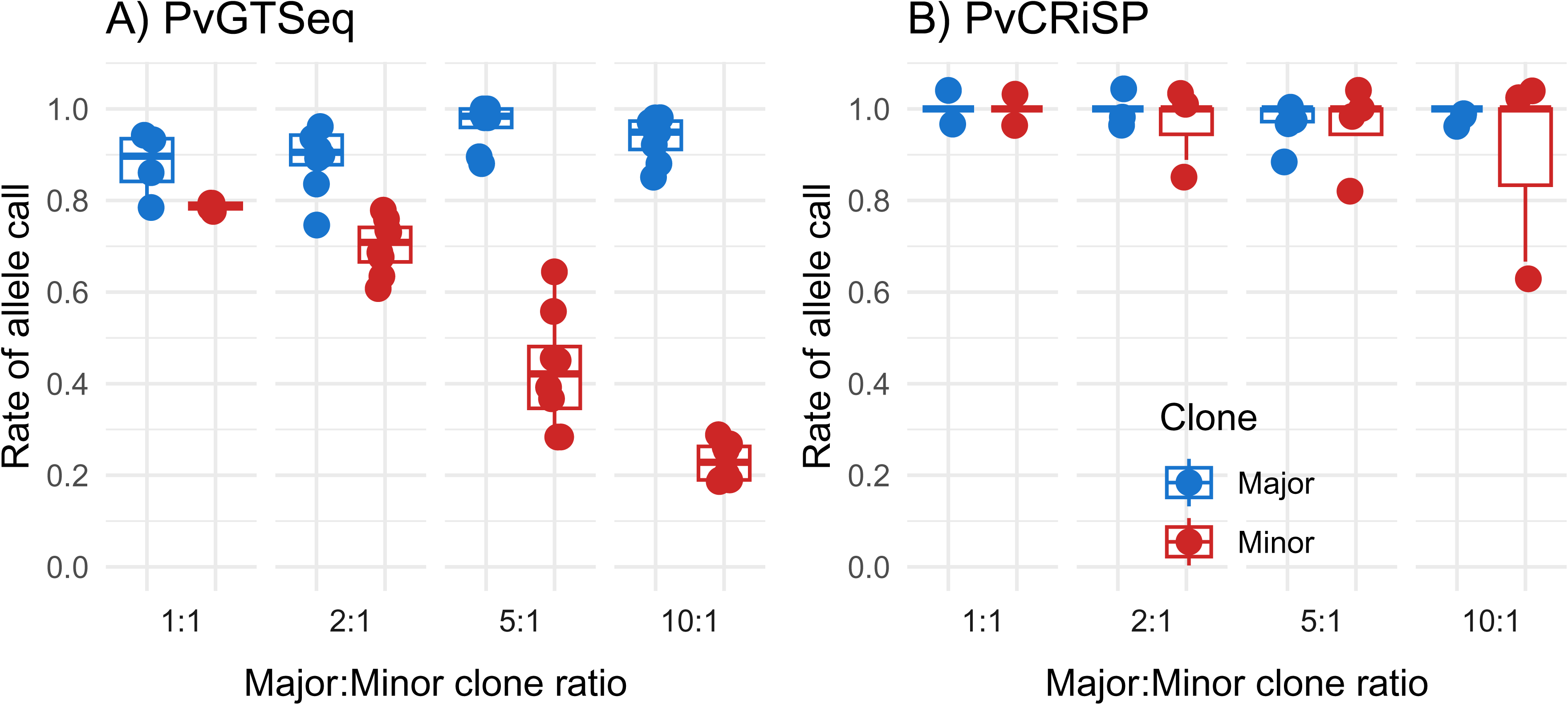
Detectability of alleles from major (blue) and minor (red) clones in mock samples using PvGTSeq (A) and PvCRiSP (B) panels. The X-axis represents the ratio of the concentration of the major clone (numerator) with respect to the minor clone (denominator) in the mock sample. The Y-axis represents the proportion of private alleles of the clone of interest detected (numerator) with respect to the total number of private alleles expected (denominator). Dots represent each clone of interest in the 28 mock samples.

### Diversity and utility of the amplicon panels

Genetic diversity varied across amplicon subsets and geographic areas at different spatial scales. Globally, from the 201 highly sensitive amplicons in PvGTSeq, the median number of polymorphic sites (SNVs and INDELs) and microhaplotypes per amplicon was 7 (IQR = 6) and 9 (IQR = 7), respectively. The nucleotide diversity (π) was 4.38e-03 (IQR = 5.13e-03) (Fig 3). Among the four PvGTSeq amplicon subsets (differentiation at three geographical scales and genes associated with antimalarial resistance), those selected for their discrimination power within Latin America and Caribbean countries showed the highest diversity (Fig S8 in S1 Text), with a median 11 microhaplotypes (IQR = 12.5), and a median nucleotide diversity of 5.61e-03 (IQR = 5.58e-03). Conversely, amplicons targeting antimalarial resistance genes showed the lowest diversity (S = 3, n.all = 4, π = 2.8e-03) (Fig 3), except for two amplicons, pvpi3k_7 and pvpi3k_13, targeting the *PI3K* gene (PVP01_1018600) which exhibited more than 30 microhaplotypes due to short tandem repeats in their sequences. While only the amplicon pvpi3k_1 was monomorphic across all global monoclonal samples, it showed polymorphism in three polyclonal samples from Cambodia, Thailand and Myanmar. Regarding PvCRiSP, it showed more than six polymorphic sites (median = 8.5, IQR = 2.75) and eight microhaplotypes per amplicon (median = 13, IQR = 17.25), with global nucleotide diversity above 3.15e-03 (median = 1.06e-02, IQR = 6.55e-03). Across the seven global regions analyzed, the median nucleotide diversity of the 169 amplicons for geographic differentiation of the PvGTSeq panel exceeded 3.01e-03 in Africa (AF), Eastern Southeast Asia (ESEA), Latin America and Caribbean (LAC), Western Southeast Asia (WSEA) and Western Asia (WAS) regions with no significant inter-regional differences (Fig 3). However, Maritime Southeast Asia (MSEA) and Oceania (OCE) showed lower nucleotide diversity (1.75e-03 and 1.7e-03, respectively), with up to 73 (36%) monomorphic amplicons in these regions. This pattern persisted across various PvGTSeq panel subsets (use groups) as well as in the PvCRiSP panel. At the country level, Brazil showed the highest nucleotide diversity (π = 5.01e-03) while Malaysia showed the lowest (π = 5.72e-04; Fig S9 in S1 Text). Indonesia and Papua New Guinea were excluded from this analysis due to insufficient monoclonal samples.

**Fig 3:**
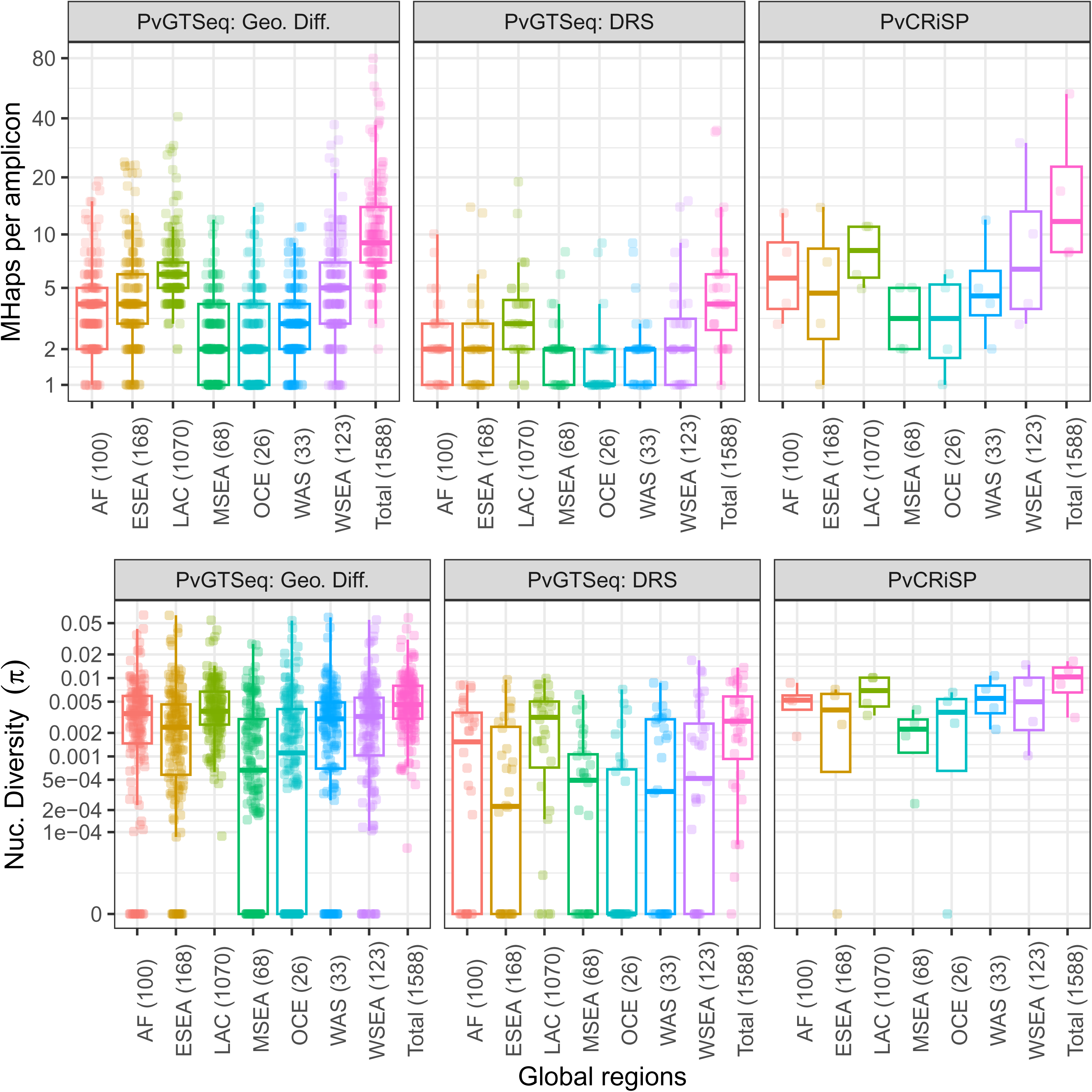
Number of microhaplotypes (MHaps) and nucleotide diversity (π) per amplicon use groups and across seven global regions: Africa (AF), Eastern Southeast Asia (ESEA), Latin America and Caribbean (LAC), Maritime Southeast Asia (MSEA), Oceania (OCE), Western Asia (WAS), and Western Southeast Asia (WSEA). Numbers within parenthesis indicate the number of monoclonal clinical samples. Each dot represents an amplicon. Use case groups were defined as 169 amplicons for geographic differentiation, 32 amplicons for drug resistance surveillance (DRS), and the four amplicons comprising PvCRiSP.

**Fig 4:**
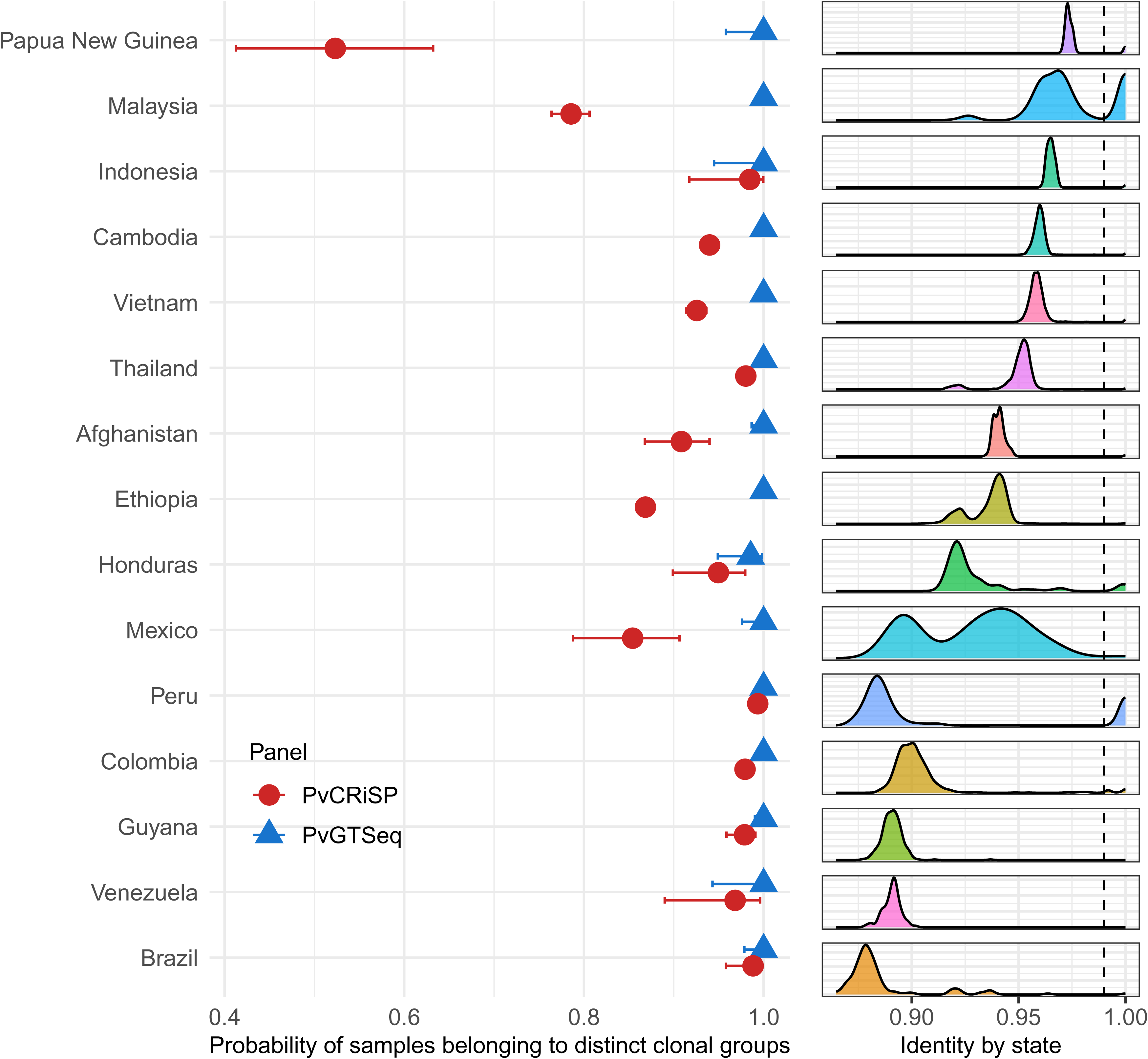
Panel discrimination power. The figure on the left shows the probability that two random samples will differ in at least one amplicon when they belong to distinct clonal groups (x-axis) across global *P. vivax* populations (y-axis) using PvGTSeq (blue) and PvCRiSP (red). The figure on the right shows the distribution of identity by state within each population measured from whole genome sequencing data and the vertical dashed line indicates the threshold used to define clonal groups (IBS ≥ 0.99).

To directly measure the utility of PvGTSeq and PvCRiSP in each country, we measured the discriminatory power of the panels at two levels: 1) differentiating parasites with different genomic sequences, and 2) differentiating parasites that belong to different clonal groups. In both cases, the discriminatory power of the panels was defined as the probability that two random samples differ from each other in at least one amplicon (which we will call D due to similarity with Simpson’s index); this means that their IBS is less than one.

Regarding the ability of the panels to differentiate parasites with different genomic sequences, for PvGTSeq, it was found that in 15 of the 16 analyzed countries, D was ≥ 0.95, but lower in Panama (0.767, 95% CI from 0.748 to 0.785) (Fig S10 in S1 Text). For PvCRiSP, D was ≥ 0.95 in 7 countries, between 0.9 and 0.95 in four countries, and below 0.9 in 5 countries (Ethiopia, Mexico, Malaysia, Panama and Papua New Guinea). In both panels, D was influenced by the distribution of IBS of the pairwise comparisons in the population (Pearson correlation p-value < 0.001, adj-R^2^ = 0.71).

Regarding the ability to differentiate parasites belonging to different clonal groups, D was 1.0 for all countries except Honduras, where it was 0.986. For PvCRiSP D was above 0.8 for all countries except Malaysia (0.786) and Papua New Guinea (0.523).

### Population structure and geographic attribution

Genetic differentiation using a PCoA and a network method revealed that genetic differences align with administrative boundaries at three geographical levels: global, intra-continental, and subnational. The global PCoA divided samples into three major groups: the first composed of LAC, the second including AF and WAS, and the third comprising ESEA, MSEA, OCE and WSEA. Each of these groups showed internal differentiation patterns (Fig 5A and Fig S11 in S1 Text). In Latin America and the Caribbean, PCoA analysis revealed that the first principal coordinate, explaining 43% of genetic differentiation, separates parasites from the Amazon region (Eastern Colombia, Guyana, and Venezuela) from those in the Colombian Pacific and Central America (Fig 5C). The second coordinate distinguished parasites from Colombia from those in Honduras and Panama. Network analysis of samples from Colombia, Honduras, and Panama further reveals distinct clonal groups specific to each sub-national region (Fig 5D and Fig S12 in S1 Text). Finally, the sequencing method used (amplicon or WGS) did not cause large batch effects (Fig S13 in S1 Text).

**Fig 5:**
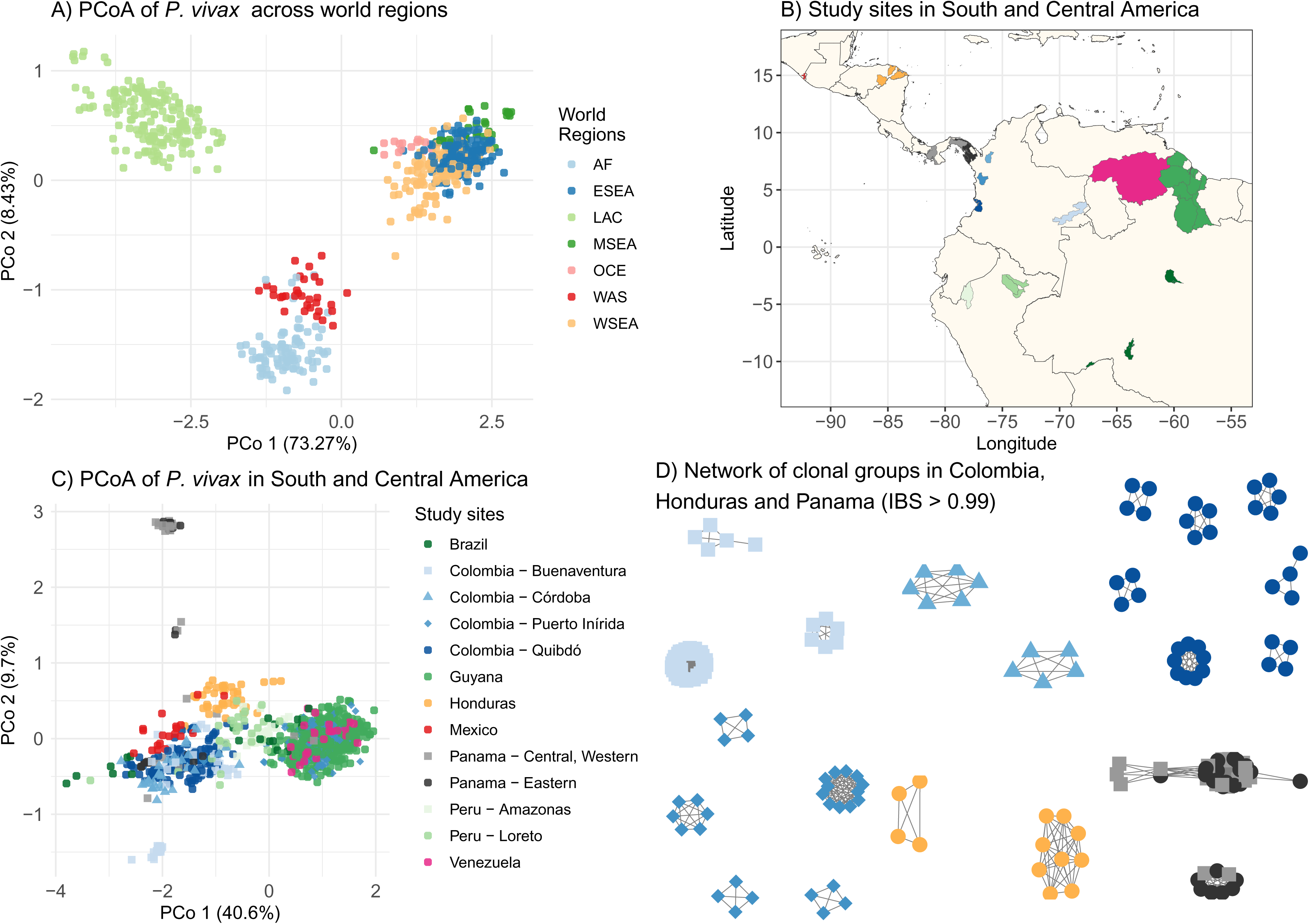
A) Principal coordinate analysis (PCoA) of *P. vivax* samples from seven global regions: Africa (AF), Eastern Southeast Asia (ESEA), Latin America and Caribbean (LAC), Maritime South-East Asia (MSEA), Oceania (OCE), Western Asia (WAS), and Western South-East Asia (WSEA). LAC samples were sequenced either by WGS or the PvGTSeq amplicon panel (shown mutually exclusively to avoid duplicates) or were obtained from the MalariaGEN Pv4 database. Genetic information for other regions was extracted from the MalariaGEN Pv4 database. Each point represents a monoclonal sample, with colors indicating geographical origin. The x- and y-axes represent principal coordinates 1 and 2, respectively. B) *P. vivax* sampling areas across South and Central America for genetic characterization using PvGTSeq. Colored regions show the administrative areas where samples were collected—first subnational level for Guyana, Mexico and Venezuela, and second subnational level for Brazil, Colombia, Honduras, and Peru. C) PCoA of *P. vivax* populations in South and Central America. Points represent monoclonal samples, with colors and shapes indicating country of origin, except for Colombia, where colors represent municipality (second subnational level). The x- and y-axes represent principal coordinates 1 and 2, respectively. D) Network graph showing genetic relationships (IBS) among samples within clonal groups (IBS > 0.99) in Colombia, Honduras, and Panama. Each node represents a monoclonal sample, with edges indicating genetic relationships exceeding 0.99 IBS. Figures B, C and D share the same color scheme.

## Discussion

We describe two new amplicon-based targeted sequencing panels for *P. vivax*, PvGTSeq and PvCRiSP. PvGTSeq is a multipurpose panel that requires sWGA pre-amplification and contains 201 highly sensitive amplicons for monitoring variants of interest in nine genes associated with antimalarial resistance (32 amplicons) and identifying population structure (169 amplicons), suitable for use in any global region but particularly optimized for Latin America. All amplicons are multiplexed in a single PCR reaction using non-proprietary reagents. With sWGA, PvGTSeq offers high sensitivity and amplifies ≥ 75% of amplicons in samples with parasitemias as low as four parasites/μL. The PvCRiSP panel consists of only four highly polymorphic amplicons and works with parasitemias ≥ 5 parasites/μL without sWGA, enabling characterization of samples with compromised DNA integrity. PvCRiSP is tailored towards estimating COI and distinguishing between infections caused by distinct clonal groups, information useful for characterizing outbreaks. We validated both panels directly with samples from five Latin American countries (Colombia, Guyana, Honduras, Panama, and Venezuela) and *in silico* across 16 countries worldwide. This information serves as a reference for potential users to evaluate the utility of these panels in their endemic areas of interest. The cost per sample of PvGTSeq library preparation and sequencing is $23.64 with 186 samples per MiSeq v2 kit 300 cycles (median of >200 reads/amplicon/sample), plus $15.99 per sample for sWGA using the Genomiphi v2 kit (manufacturer). In contrast, PvCRiSP costs $19.04 per sample due to its simplified design (S9 Table).

These panels contribute to the expanding repertoire of targeted next-generation sequencing approaches for *P. vivax* characterization, all of which have strengths and weaknesses for various use cases. The most common applications of targeted sequencing include analyzing population structure, determining geographic origin, tracking resistance variants, measuring COI, determining pairwise relatedness, and distinguishing between new infections, relapses, and recrudescences. (13,14,16–20,64) Spatial resolution depends on the genomic dataset used for target selection and the criteria applied to maximize resolution per area of interest. While MalariaGEN Pv4 data has enabled new panel development, some regions remain underrepresented— particularly Latin America—causing variable panel performance across regions.

PvAmpliSeq, the first validated amplicon sequencing panel, was designed to identify patterns of population structure between global regions and within Vietnam (14) but showed poor discrimination in South America, leading to a second version with 41 additional SNPs for Peruvian samples. A recent MIP-based panel targeting ∼1200 SNPs tested with Peruvian samples offers deep profiling of samples but struggles with parasitemias below 100 parasites/μL. (19) While previous panels were designed to characterize SNPs, two additional panels were developed to amplify microhaplotypes (multiple polymorphic sites within a single amplicon), though these had constraints with low parasitemia samples. (17,64) In creating our new PvGTSeq and PvCRiSP panels, we incorporated extensive WGS data from MalariaGEN Pv4 (genomes from Brazil, Colombia, Mexico, and Peru) and we additionally leveraged in-house WGS representing Colombia, Guyana, Peru, and Venezuela. Then we validated both panels with samples from Colombia, Guyana, Honduras, Panama and Venezuela. Additionally, we coupled PvGTSeq library preparation with sWGA. As a result, these two panels are the most sensitive and extensively evaluated in the Americas.

Distinguishing between new infections, relapses, and recrudescences requires high sensitivity to detect alleles from minority clones in polyclonal infections. In this context, panels with large numbers of multiplexed amplicons present a significant disadvantage. (17,20) Our results show that while PvGTSeq can correctly identify whether an infection is monoclonal or polyclonal, alleles from the minority clone may remain undetected up to 70% of the time when the proportion between clones is 10:1. Although reducing the number of samples per sequencing run could increase read depth and minimize allele loss from minority clones, this approach would raise costs, making this technology prohibitively expensive. This challenge is effectively addressed by using panels with fewer amplicons, such as PvCRiSP, which offers high sensitivity, efficiently detects all alleles present in polyclonal samples without requiring sWGA, and offers low procedural complexity.

As amplicon sequencing becomes more widespread for generating genetic data for malaria parasites and other pathogens, there is a need for versatile analysis tools that work with any amplicon panel. Here we used a DADA2-based analysis pipeline for denoising PCR errors and achieving sensitive minor allele detection in polyclonal infections. We validated our error-filtering approach using technical replicates at three levels: within runs, between runs, and against WGS. This multi-level validation achieved lower error rates (3.85e-4% within sequencing runs and 2.86e-3% when comparing amplicon sequencing to WGS) than comparable studies (from 0.008% to 0.02%) (10) and demonstrated that INDELs do not reduce reproducibility in amplicon sequencing.

This panel-agnostic pipeline is available on GitHub and embedded in a Terra workspace—a cloud-based platform designed for users with limited bioinformatics expertise (S2 Text). A comprehensive description of this analysis pipeline and all its functionalities will be presented in a subsequent publication.

Despite their significant advantages, both PvGTSeq and PvCRiSP have limitations. These include ascertainment bias inherent in the design of any targeted sequencing panel, limited overlap with existing panels, and PvGTSeq allelic dropout in polyclonal samples with skewed strain ratios. However, despite these limitations, PCoA and network analysis confirm that PvGTSeq can identify population structure on a global scale as well as detect clonal expansions at subnational levels in Colombia and Panama, where persistent clonal groups have been previously documented using WGS. (65,66)

The results presented here demonstrate that PvGTSeq and PvCRiSP are two highly sensitive panels that allow genetic characterization of infections above five parasites/μL. Through analysis of samples from five endemic countries in Latin America and the Caribbean, we demonstrate that PvGTSeq enables monitoring of candidate antimalarial resistance variants and identification of population structure at different geographic scales. Meanwhile, PvCRiSP provides reliable estimation of COI and identifies cases of clonal transmission even in samples with compromised DNA integrity. This information will serve as a reference for describing demographic changes in the *P. vivax* population, particularly in Latin America and the Caribbean—a region that in recent years has faced significant challenges for malaria control and elimination.

## Supporting information

S3 Text: PvCRiSP protocol

S2 Text: PvGTSeq protocol

## Data Availability

The sequence data generated by this study has been deposited in the NCBI Sequence Read Archive (https://www.ncbi.nlm.nih.gov/sra) under BioProject PRJNA1092690. Data preparation scripts are available at https://github.com/Paulonvnv/PvGTSeq_PvCRiSP_paper. All other data are included in the manuscript and supporting information.

https://github.com/Paulonvnv/PvGTSeq_PvCRiSP_paper

## Acknowledgements

This work was supported, in whole or in part, by the Bill & Melinda Gates Foundation INV-043618. This project has also been funded in whole or in part with Federal funds from the National Institute of Allergy and Infectious Diseases, NIH, Department of Health and Human Services, under Grant U19Al110818 to the Broad Institute. We thank the participants who contributed blood samples to the study, as well as the technicians who collected and processed the samples.

## Author contributions

**Conceptualization**: Paulo C. Manrique-Valverde, Chloe M. Hasund, Sarah Auburn, Daniel E. Neafsey.

**Data Curation**: Katrina A. Kelley, Margaret Laws, Raphael Brosula, Ana M. Santamaria, Stella M. Chenet, Reza Niles-Robin, Myriam Arévalo-Herrera, Sócrates Herrera, Vladimir Corredor, Justin Lana, David Forero-Peña, Gustavo Fontecha, Jose Calzada, Nicanor Obaldia.

**Formal Analysis**: Paulo C. Manrique-Valverde, Chloe M. Hasund.

**Funding Acquisition:** Sarah Auburn, Daniel E. Neafsey.

**Investigation and Methodology:** Paulo C. Manrique-Valverde, Katrina A. Kelley, Chloe M. Hasund.

**Project Administration and Resources:** Margaret Laws, Katrina A. Kelley.

**Software:** Paulo C. Manrique-Valverde, Jorge-Eduardo Amaya-Romero, Philipp Schwabl, Raphael Brosula, Angela M. Early.

**Supervision:** Daniel E. Neafsey.

**Validation:** Paulo C. Manrique-Valverde, Chloe M. Hasund, Katrina A. Kelley.

**Visualization:** Paulo C. Manrique-Valverde.

**Writing – Original Draft Preparation:** Paulo C. Manrique-Valverde, Chloe M. Hasund, Daniel E. Neafsey.

**Writing – Review & Editing:** All authors reviewed the manuscript.

## Supporting information

**S1 Text: Supporting figures**. **Fig S1:** Flowchart of the selection, optimization and validation process for PvGTSeq and PvCRiSP. Each stage details the number of amplicons (segments or primer pairs) selected, the samples or genomic data used for the analysis, and the evaluation criteria applied.

**S1 Text: Supporting figures**. **Fig S2:** Examples of Pseudo-cigar representation of polymorphisms observed from multiple alignment of amplicon microhaplotypes with their reference sequence. In our analysis pipeline, after dada2 denoising of sequencing errors, all microhaplotypes are aligned to the reference sequence using MUSCLE (Multiple Sequence Comparison by Log-Expectation). Polymorphisms are summarized in Pseudo-cigar format following these rules: 1) All variants are annotated in ascending order. 2) For SNVs, we annotated the reference position followed by the substitute nucleotide. 3) For deletions, we annotated the starting position, "D=", then all deleted nucleotides (e.g., 23D=TG). 4) For insertions, we annotated the position before insertion, "I=", then the nucleotide at that position followed by inserted nucleotides. 5) If the nucleotide before an insertion is an SNV, we include this SNV within the insertion notation to avoid position duplication (see MHAP05). 6) When a deletion is followed by an insertion, the insertion is annotated first to maintain ascending order (see MHAP03 and MHAP04). 7) For insertions before position 1, we use position 0 and only annotate inserted nucleotides (see MHAP05-MHAP07). 8) If both reference and microhaplotype have deletions, but the microhaplotype deletion extends beyond the reference deletion, we consider it a single deletion (see MHAP06 and MHAP07). 9) If both, the reference and the microhaplotype, have deletions, but the reference deletion extends beyond the microhaplotype deletion, we consider it a single insertion (see MHAP05). 10) Microhaplotypes that are identical to the reference sequence are annotated using a period symbol "." (see MHAP01).

**S1 Text: Supporting figures**. **Fig S3:** Bar plot showing the prevalence of haplotypes of genes that carry mutations associated with antimalarial resistance. y-axis shows the frequency in each population, x-axis shows the country in which the sample was collected, horizontal sections correspond to the analyzed gene, and vertical sections represent each world region.

**S1 Text: Supporting figures**. **Fig S4:** A) Scatter plot of the median read depth and the percentage of amplified samples of each amplicon on each of three populations: Colombia (Green), Guyana (Red) and Venezuela (Blue). Dots represent each of the 249 amplicons in PvGTSeq while the letters represent the 4 amplicons in PvCRiSP (CG2_releated, RIPR, VSP11, and PIGM). B) Distribution of amplification rate of the amplicons in PvGTSeq and PvCRiSP in the three countries, as indicated by color.

Numbers in figure B indicate the number of amplicons in each population that are in the 1st, 2nd, 3rd, and 4th quantile. C) Distribution of limit of detection (LOD) of each individual amplicon in PvGTSeq and PvCRiSP. D) Scatter plot of amplification rate (x- axis) and LOD (y-axis) of each individual amplicon (dots or letters) in three populations.

**S1 Text: Supporting figures**. **Fig S5:** Contribution of each amplicon and the type of polymorphism to the discrepancies within and between PvGTSeq sequencing runs, and between PvGTSeq with respect to WGS. X-axis shows the number of times an amplicon (y-axis) showed a discrepancy between technical replicates in any of the 3 experiments (Vertical panels): Within and between sequencing runs of PvGTSeq, and between PvGTSeq and WGS. Colors indicate if the discrepancy in the amplicon was due to a single nucleotide variant (SNV, in gold), an insertion or deletion (INDEL, in red) or any of both (in blue).

**S1 Text: Supporting figures**. **Fig S6:** The top panels show the fraction of heterozygous loci detected relative to the ratio of major and minor clones in mock samples using PvGTSeq (A) and PvCRiSP (B). The bottom panels illustrate the correlation between the read depth ratio of major and minor clones for PvGTSeq (C) and PvCRiSP (D) compared to their DNA concentration ratios. MS1 and MS2 represent the two biological replicates, and shapes of the dots (circles and triangles) represent the 2 technical replicates.

**S1 Text: Supporting figures**. **Fig S7:** Read depth of major and minor clones within mock samples. Each dot represents a private microhaplotype from major (blue) and minor (red) clones using the 201 amplicon from PvGTSeq. Horizontal panels represent each of the two combinations of mock samples (MS1 and MS2) and their technical replicates (R1 and R2), while the vertical panels represent the different ratios at which mock samples were generated.

**S1 Text: Supporting figures**. **Fig S8:** Number of polymorphic sites, microhaplotypes (MHaps)alleles and nucleotide diversity (π) per amplicon use groups and across seven global regions: Africa (AF), Eastern South-East Asia (ESEA), Latin America and Caribbean (LAC), Maritime South-East Asia (MSEA), Oceania (OCE), Western Asia (WAS), and Western South-East Asia (WSEA). Numbers within parenthesis indicate the number of monoclonal clinical samples. Each dot represents an amplicon. Each dot represents an amplicon. Use case groups were defined as 81 amplicons for geographic differentiation between world regions (WR), 49 amplicons for geographic differentiation between countries in LAC (LACbc), 35 amplicons for geographic differentiation within countries in LAC (LACwc), 32 amplicons for drug resistance surveillance (DRS), and the four amplicons comprising PvCRiSP. Because there are amplicons for geographic differentiation that belong to multiple groups, to avoid duplication we assigned the amplicon to the highest geographical scale.

**S1 Text: Supporting figures**. **Fig S9:** Number of polymorphic sites, microhaplotypes (MHaps) and nucleotide diversity (π) per amplicon use-case groups and across 13 countries: Ethiopia (ETH), Cambodia (KHM), Vietnam (VNM), Brazil (BRA), Colombia (COL), Guyana (GUY), Honduras (HND), Panama (PAN), Peru (PER), Venezuela (VEN), Malaysia (MYS), Afghanistan (AFG) and Thailand (THA). Each dot represents an amplicon. Use case-groups were defined as 169 amplicons for geographic differentiation, 32 amplicons for drug resistance surveillance (DRS), and the four amplicons conforming PvCRiSP.

**S1 Text: Supporting figures**. **Fig S10:** Panel discrimination power. The figure on the left shows the probability that two random samples differ from each other in at least one amplicon (x-axis) across global *P. vivax* populations (y-axis) using PvGTSeq (blue) and PvCRiSP (red). Figure on the right shows the distribution of identity by state within each population measured from whole genome sequencing data.

**S1 Text: Supporting figures**. **Fig S11:** Principal coordinate analysis between Africa (AF) and Western Asia (WAS) in panel A, and between Eastern South-East Asia (ESEA), Maritime South-East Asia (MSEA), Oceania (OCE), and Western South-East Asia (WSEA) in panel B.

**S1 Text: Supporting figures**. **Fig S12:** Network graph showing genetic relationships (IBS) among samples from Colombia, Honduras, Mexico and Panama. Each node represents a monoclonal sample, with edges indicating genetic relationships exceeding 0.6 IBS. Figures B, C and D share the same color scheme.

**S1 Text: Supporting figures**. **Fig S13:** Principal coordinate analysis of samples from Guyana generated through PvGTSeq (light blue dots) and WGS (blue dots).

**S2 Text: PvGTSeq protocol. S3 Text: PvCRiSP protocol.**

**S4 Table: List of amplicons included in the PvGTSeq and PvCRisP panel.** (XSLX)

**S5 Table: Prevalence by country of haplotypes of genes that carry mutations associated with antimalarial resistance.** (XSLX)

**S6 Table: LOD, amplification rate and read depth of each amplicon in PvGTSeq by country.** (XSLX)

**S7 Table: LOD of PvGTSeq and PvCRiSP by country.** (XSLX)

**S8 Table: LOD, amplification rate and read depth of each amplicon in PvCRiSP by country.** (XSLX)

**S9 Table: Cost breakdown of PvGTSeq and PvCRiSP.** (XSLX)

