## Supplementary material for "PvGTSeq and PvCRiSP: two amplicon-based targeted sequencing panels for *Plasmodium vivax*": S1 Text: Supporting figures: S1_Text.docx

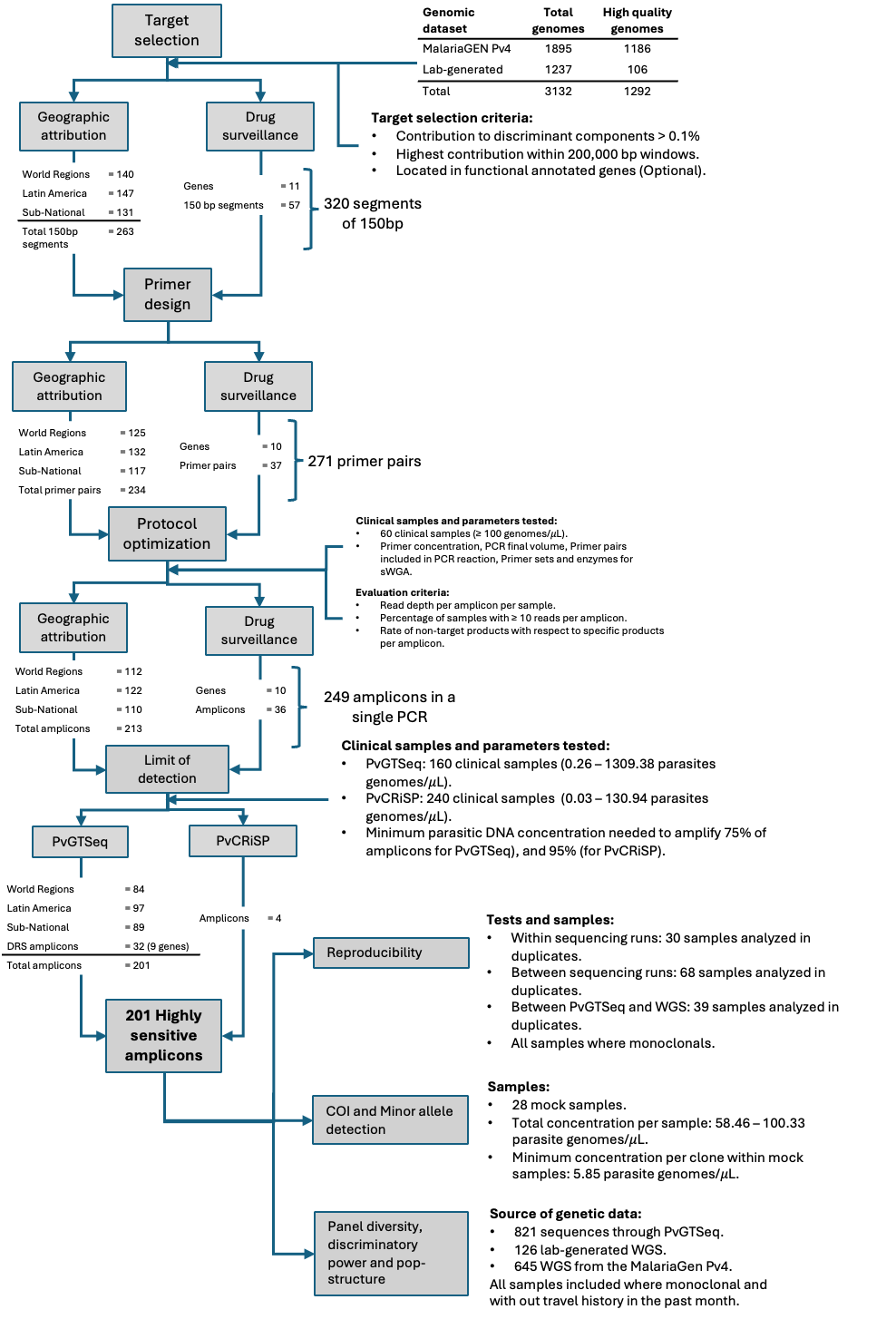

**Fig S1:** Flowchart of the selection, optimization and validation process for PvGTSeq and PvCRiSP. Each stage details the number of amplicons (segments or primer pairs) selected, the samples or genomic data used for the analysis, and the evaluation criteria applied.

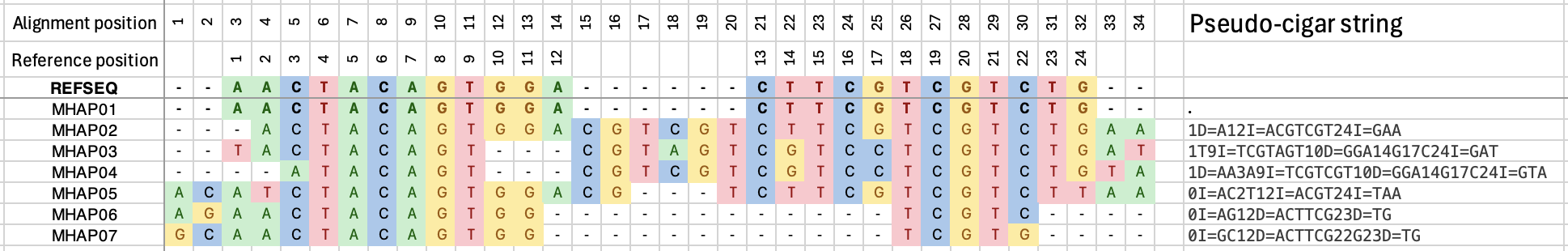

**Fig S2:** Examples of Pseudo-cigar representation of polymorphisms observed from multiple alignment of amplicon microhaplotypes with their reference sequence. In our analysis pipeline, after dada2 denoising of sequencing errors, all microhaplotypes are aligned to the reference sequence using MUSCLE (Multiple Sequence Comparison by Log-Expectation). Polymorphisms are summarized in Pseudo-cigar format following these rules: 1) All variants are annotated in ascending order. 2) For SNVs, we annotated the reference position followed by the substitute nucleotide. 3) For deletions, we annotated the starting position, "D=", then all deleted nucleotides (e.g., 23D=TG). 4) For insertions, we annotated the position before insertion, "I=", then the nucleotide at that position followed by inserted nucleotides. 5) If the nucleotide before an insertion is an SNV, we include this SNV within the insertion notation to avoid position duplication (see MHAP05). 6) When a deletion is followed by an insertion, the insertion is annotated first to maintain ascending order (see MHAP03 and MHAP04). 7) For insertions before position 1, we use position 0 and only annotate inserted nucleotides (see MHAP05-MHAP07). 8) If both reference and microhaplotype have deletions, but the microhaplotype deletion extends beyond the reference deletion, we consider it a single deletion (see MHAP06 and MHAP07). 9) If both, the reference and the microhaplotype, have deletions, but the reference deletion extends beyond the microhaplotype deletion, we consider it a single insertion (see MHAP05). 10) Microhaplotypes that are identical to the reference sequence are annotated using a period symbol "." (see MHAP01).

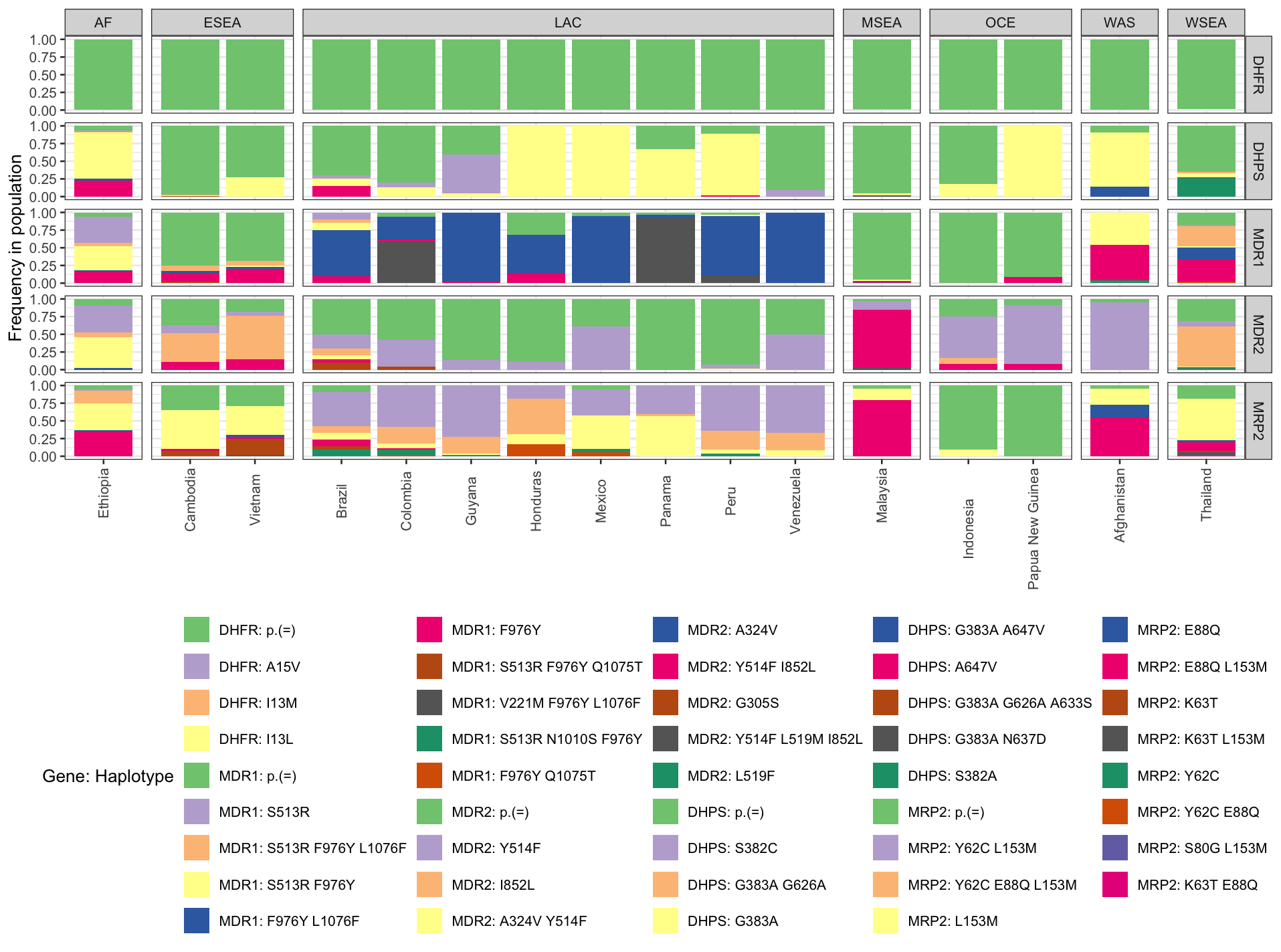

**Fig S3:** Bar plot showing the prevalence of haplotypes of genes that carry mutations associated with antimalarial resistance. y-axis shows the frequency in each population, x-axis shows the country in which the sample was collected, horizontal sections correspond to the analyzed gene, and vertical sections represent each world region.

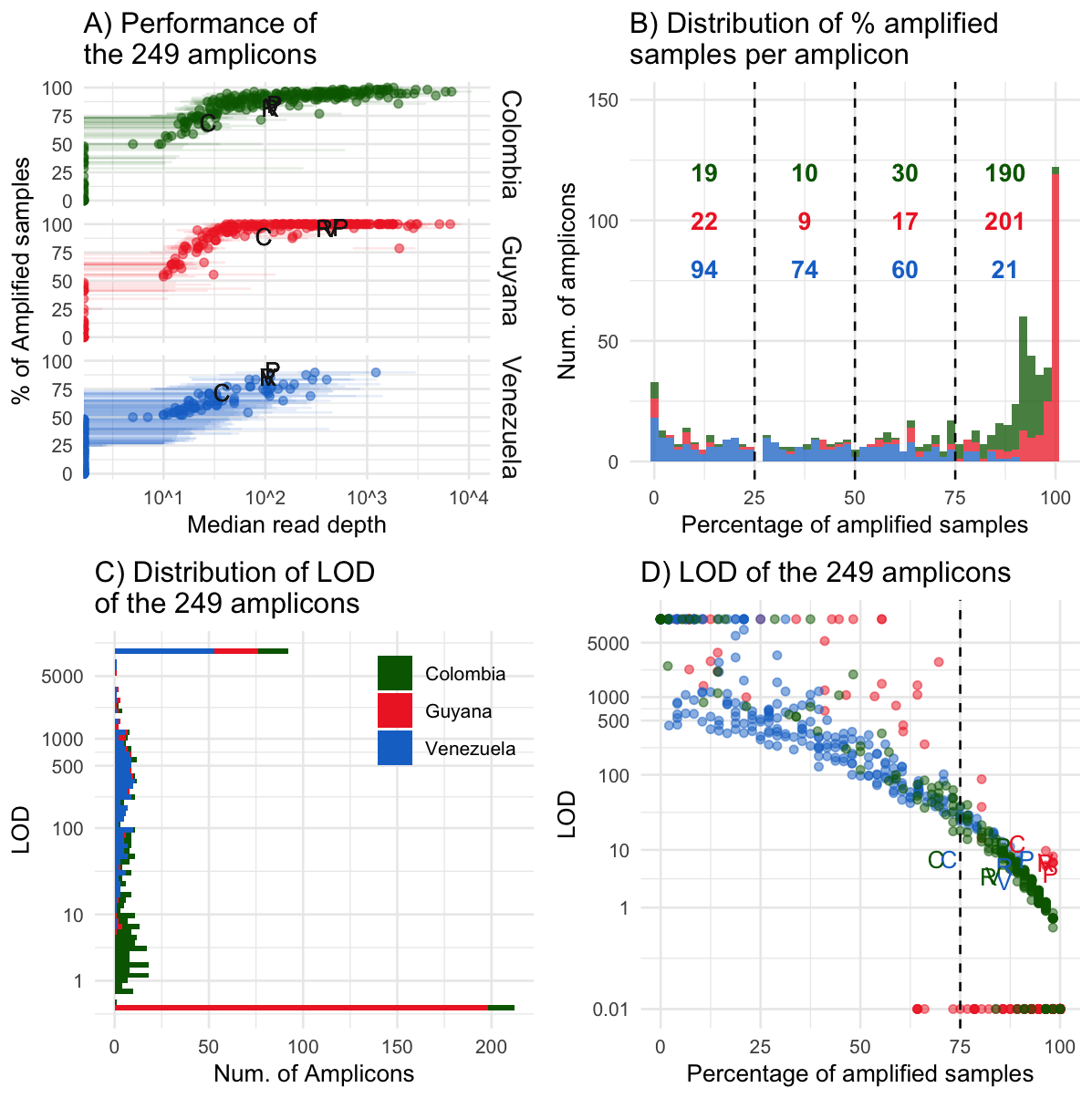

**Fig S4:** A) Scatter plot of the median read depth and the percentage of amplified samples of each amplicon on each of three populations: Colombia (Green), Guyana (Red) and Venezuela (Blue). Dots represent each of the 249 amplicons in PvGTSeq while the letters represent the 4 amplicons in PvCRiSP (CG2_releated, RIPR, VSP11, and PIGM). B) Distribution of amplification rate of the amplicons in PvGTSeq and PvCRiSP in the three countries, as indicated by color. Numbers in figure B indicate the number of amplicons in each population that are in the 1st, 2nd, 3rd, and 4th quantile. C) Distribution of limit of detection (LOD) of each individual amplicon in PvGTSeq and PvCRiSP. D) Scatter plot of amplification rate (x-axis) and LOD (y-axis) of each individual amplicon (dots or letters) in three populations.

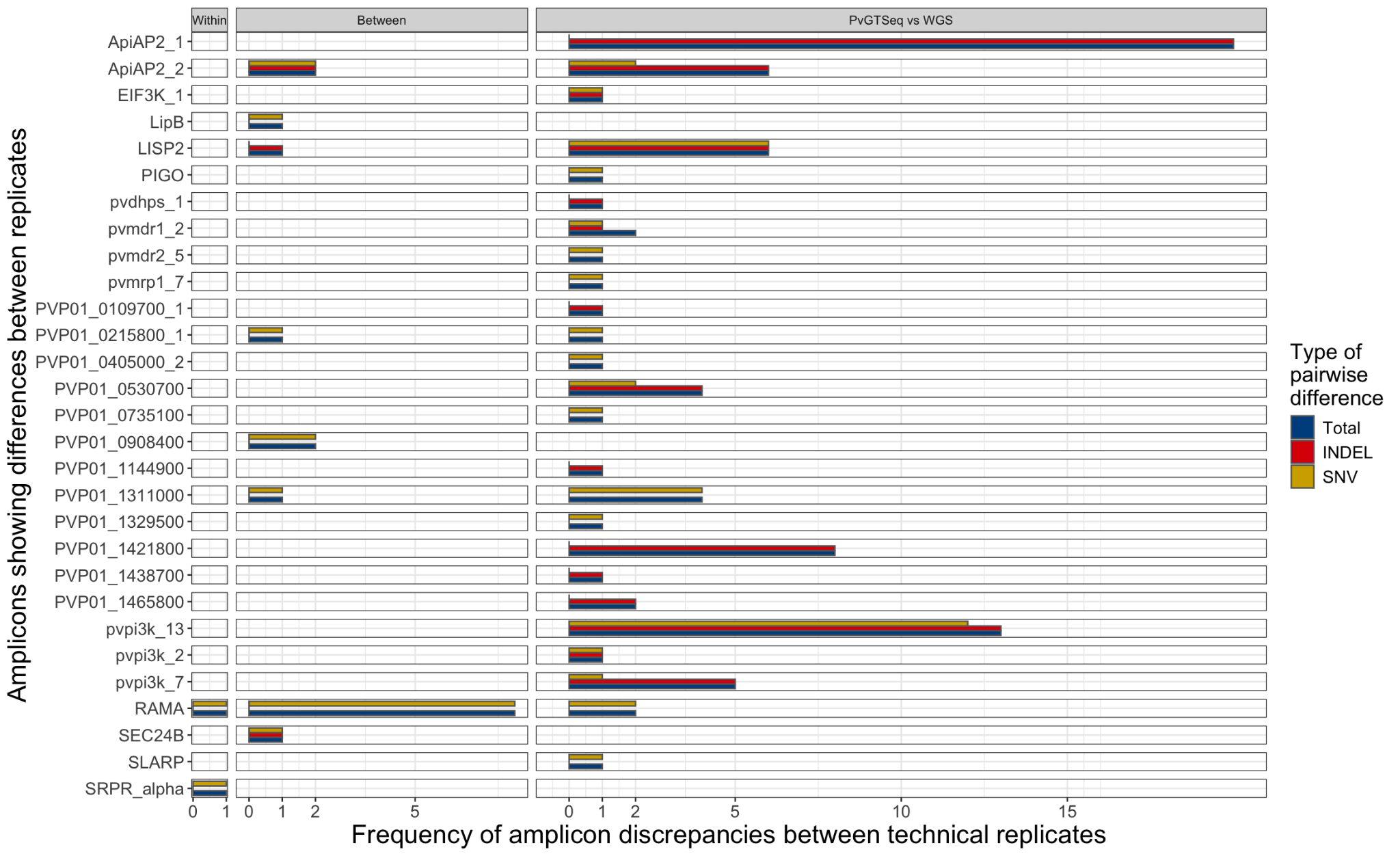

**Fig S5:** Contribution of each amplicon and the type of polymorphism to the discrepancies within and between PvGTSeq sequencing runs, and between PvGTSeq with respect to WGS. X-axis shows the number of times an amplicon (y-axis) showed a discrepancy between technical replicates in any of the 3 experiments (Vertical panels): Within and between sequencing runs of PvGTSeq, and between PvGTSeq and WGS. Colors indicate if the discrepancy in the amplicon was due to a single nucleotide variant (SNV, in gold), an insertion or deletion (INDEL, in red) or any of both (in blue).

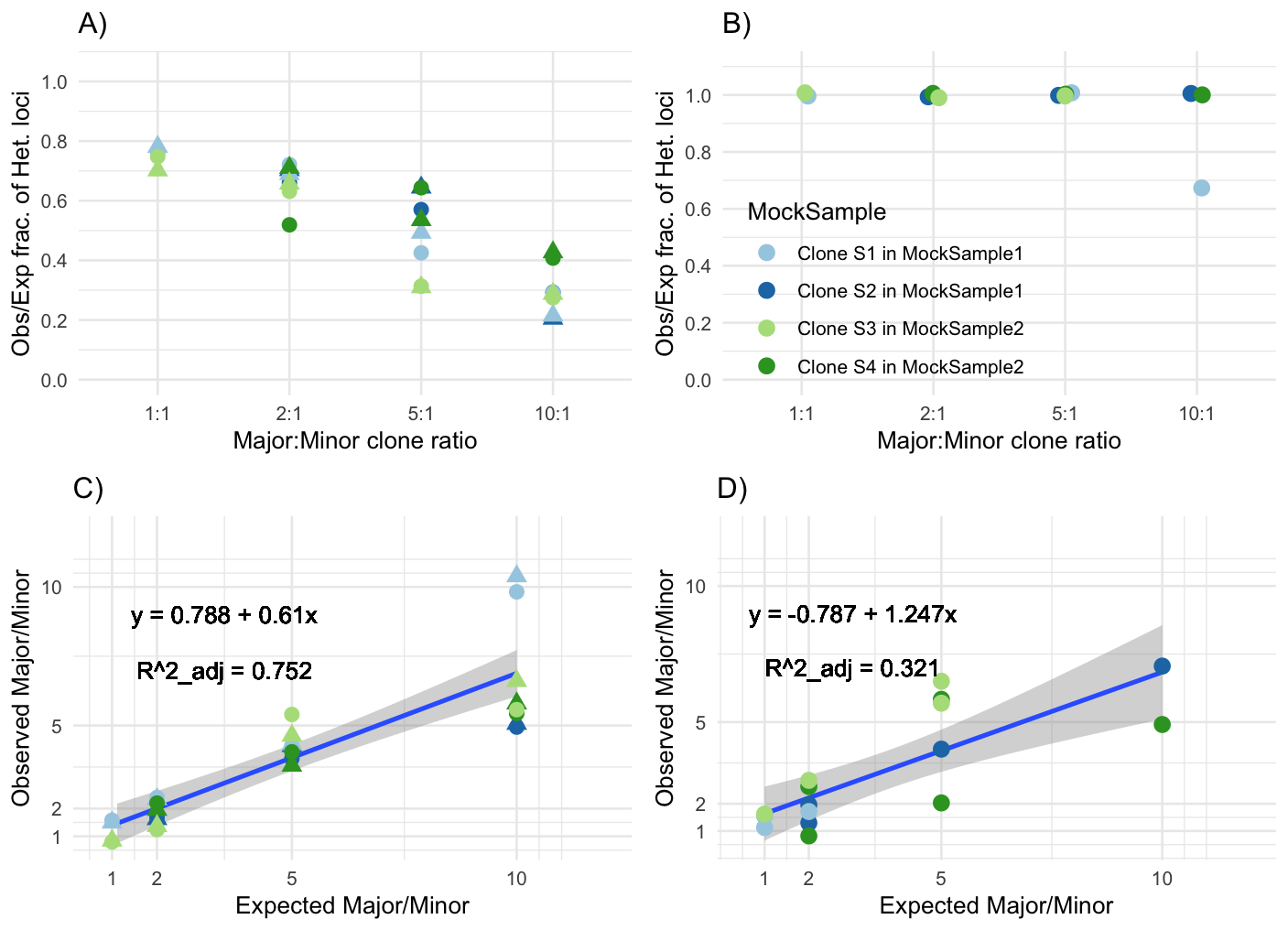

**Fig S6:**  The top panels show the fraction of heterozygous loci detected relative to the ratio of major and minor clones in mock samples using PvGTSeq (A) and PvCRiSP (B). The bottom panels illustrate the correlation between the read depth ratio of major and minor clones for PvGTSeq (C) and PvCRiSP (D) compared to their DNA concentration ratios. MS1 and MS2 represent the two biological replicates, and shapes of the dots (circles and triangles) represent the 2 technical replicates.

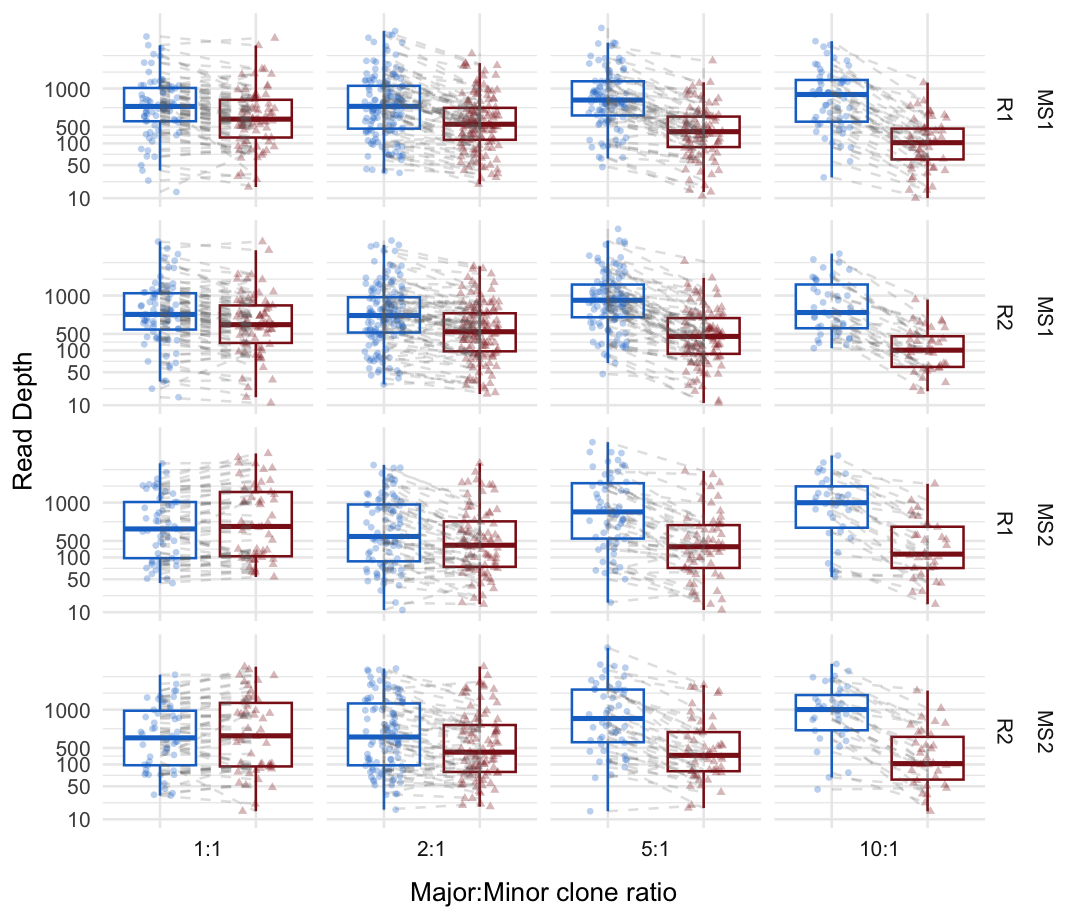

**Fig S7:**  Read depth of major and minor clones within mock samples. Each dot represents a private microhaplotype from major (blue) and minor (red) clones using the 201 amplicon from PvGTSeq. Horizontal panels represent each of the two combinations of mock samples (MS1 and MS2) and their technical replicates (R1 and R2), while the vertical panels represent the different ratios at which mock samples were generated.

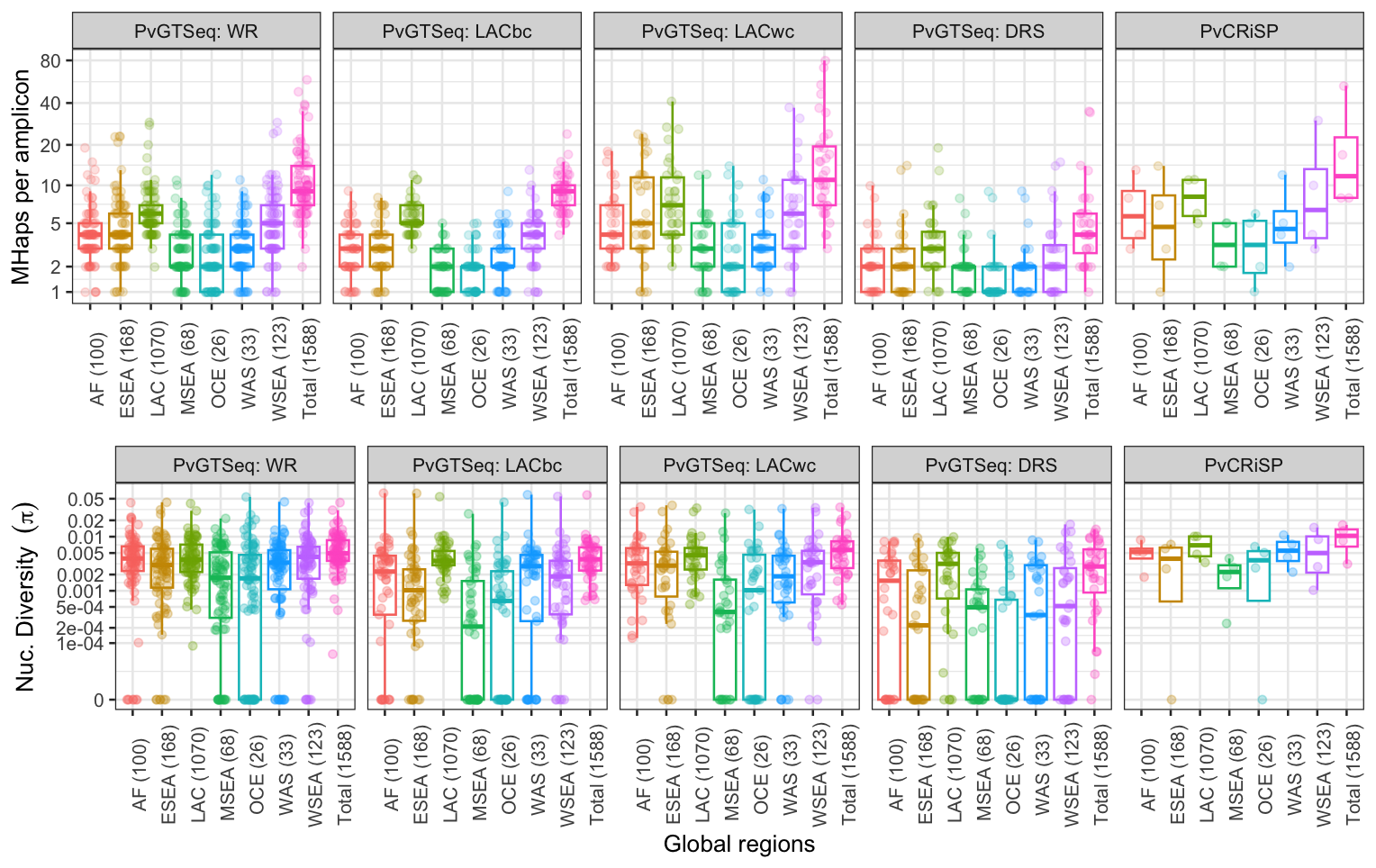

**Fig S8:** Number of polymorphic sites, microhaplotypes (MHaps)alleles and nucleotide diversity (π) per amplicon use groups and across seven global regions: Africa (AF), Eastern South-East Asia (ESEA), Latin America and Caribbean (LAC), Maritime South-East Asia (MSEA), Oceania (OCE), Western Asia (WAS), and Western South-East Asia (WSEA). Numbers within parenthesis indicate the number of monoclonal clinical samples. Each dot represents an amplicon. Each dot represents an amplicon. Use case groups were defined as 81 amplicons for geographic differentiation between world regions (WR), 49 amplicons for geographic differentiation between countries in LAC (LACbc), 35 amplicons for geographic differentiation within countries in LAC (LACwc), 32 amplicons for drug resistance surveillance (DRS), and the four amplicons comprising PvCRiSP. Because there are amplicons for geographic differentiation that belong to multiple groups, to avoid duplication we assigned the amplicon to the highest geographical scale.

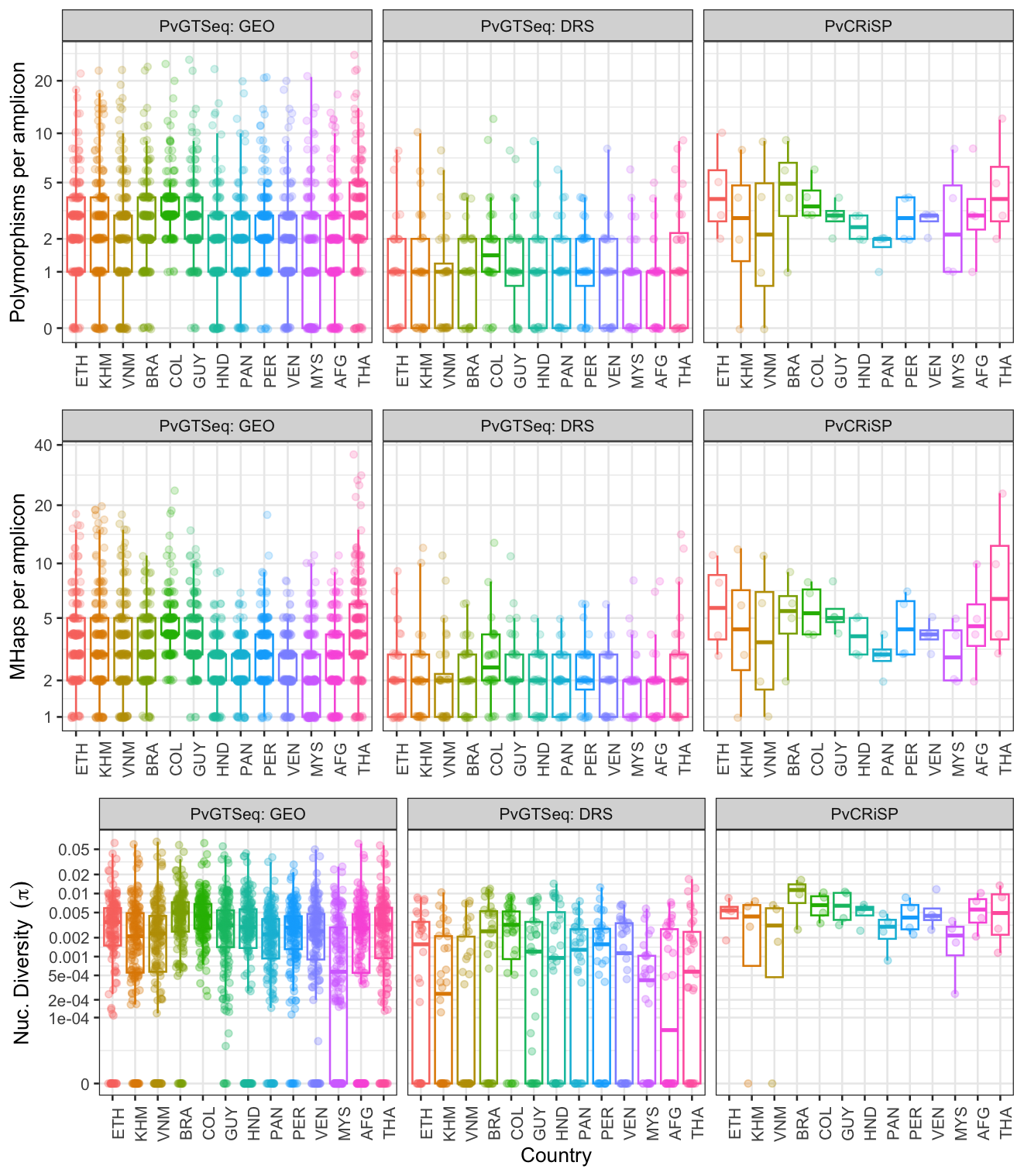

**Fig S9:** Number of polymorphic sites, microhaplotypes (MHaps) and nucleotide diversity (π) per amplicon use-case groups and across 13 countries: Ethiopia (ETH), Cambodia (KHM), Vietnam (VNM), Brazil (BRA), Colombia (COL), Guyana (GUY), Honduras (HND), Panama (PAN), Peru (PER), Venezuela (VEN), Malaysia (MYS), Afghanistan (AFG) and Thailand (THA). Each dot represents an amplicon. Use case-groups were defined as 169 amplicons for geographic differentiation, 32 amplicons for drug resistance surveillance (DRS), and the four amplicons conforming PvCRiSP.

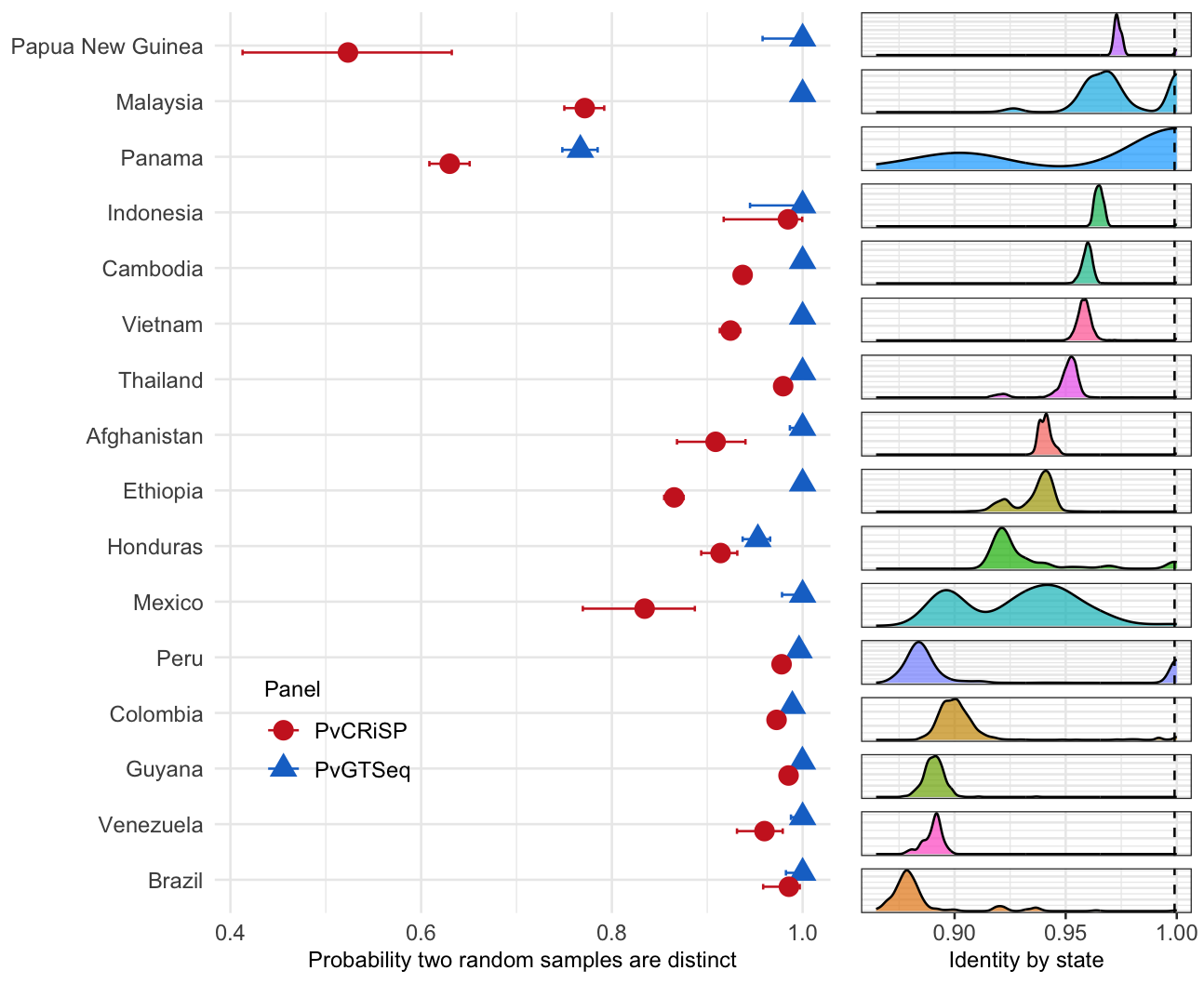

**Fig S10:** Panel discrimination power. The figure on the left shows the probability that two random samples differ from each other in at least one amplicon (x-axis) across global *P. vivax* populations (y-axis) using PvGTSeq (blue) and PvCRiSP (red). Figure on the right shows the distribution of identity by state within each population measured from whole genome sequencing data.

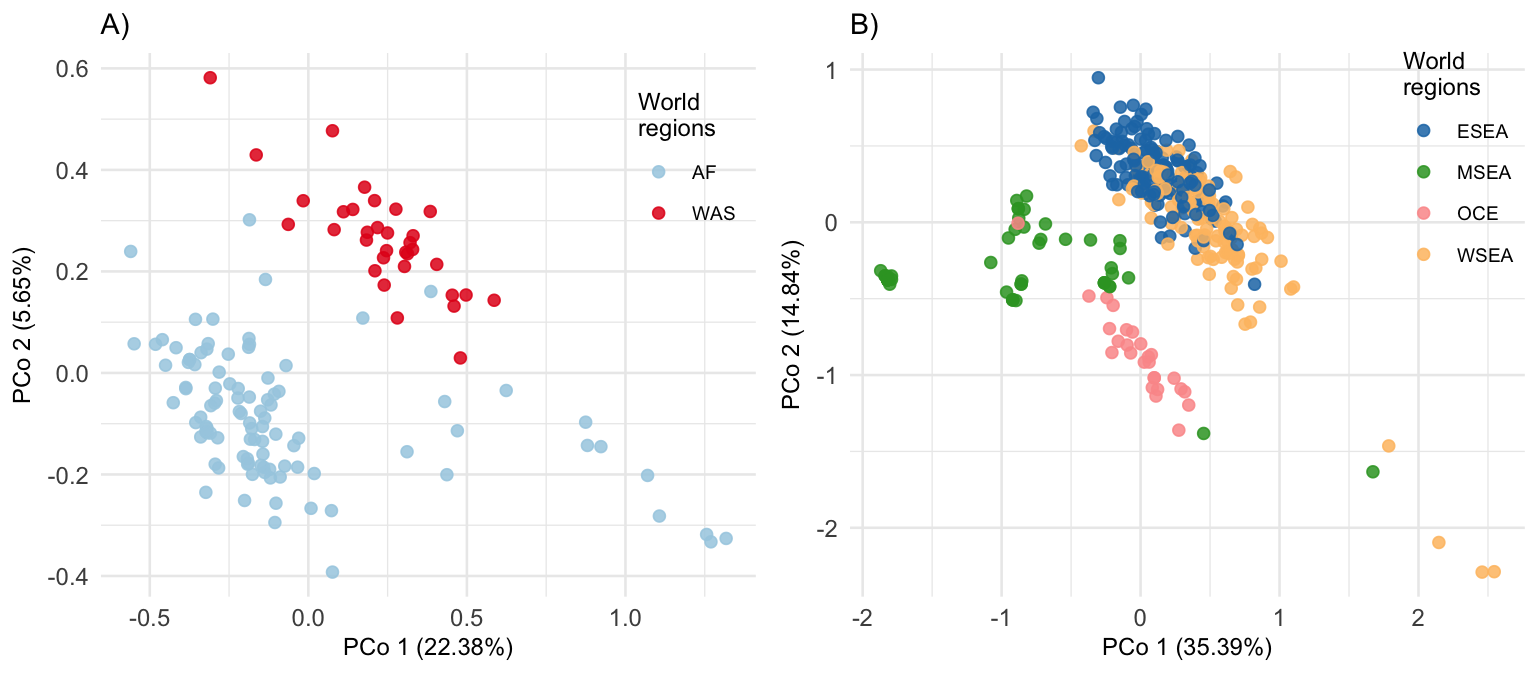

**Fig S11:** Principal coordinate analysis between Africa (AF) and Western Asia (WAS) in panel A, and between Eastern South-East Asia (ESEA), Maritime South-East Asia (MSEA), Oceania (OCE), and Western South-East Asia (WSEA) in panel B.

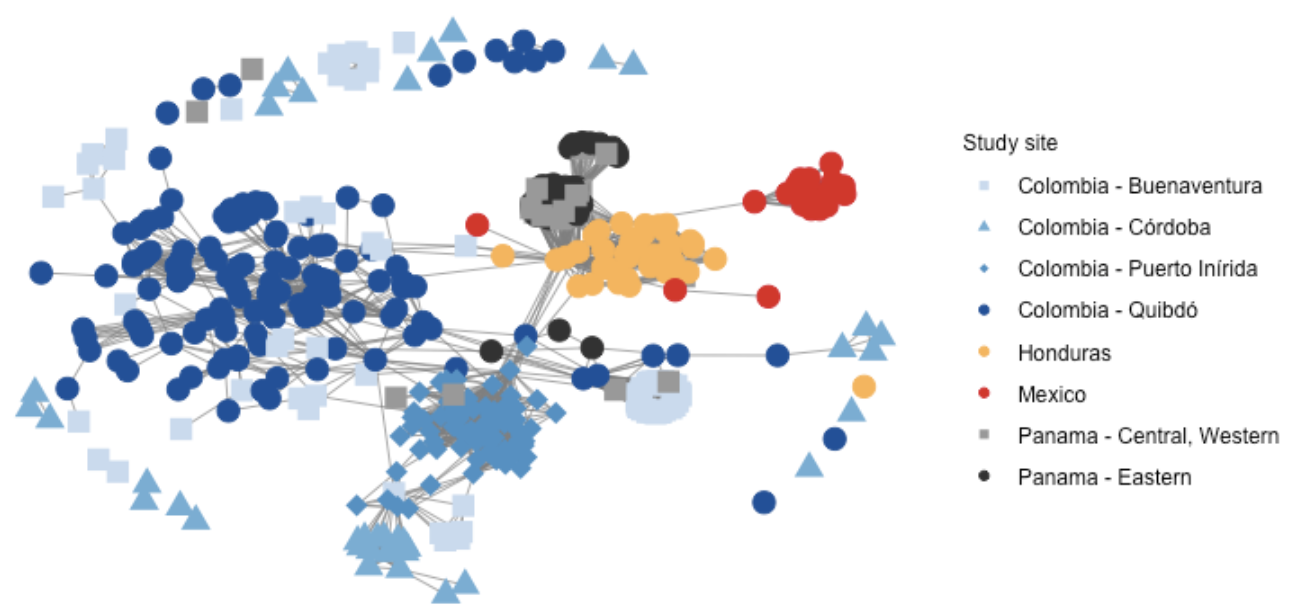

**Fig S12:** Network graph showing genetic relationships (IBS) among samples from Colombia, Honduras, Mexico and Panama. Each node represents a monoclonal sample, with edges indicating genetic relationships exceeding 0.6 IBS. Figures B, C and D share the same color scheme.

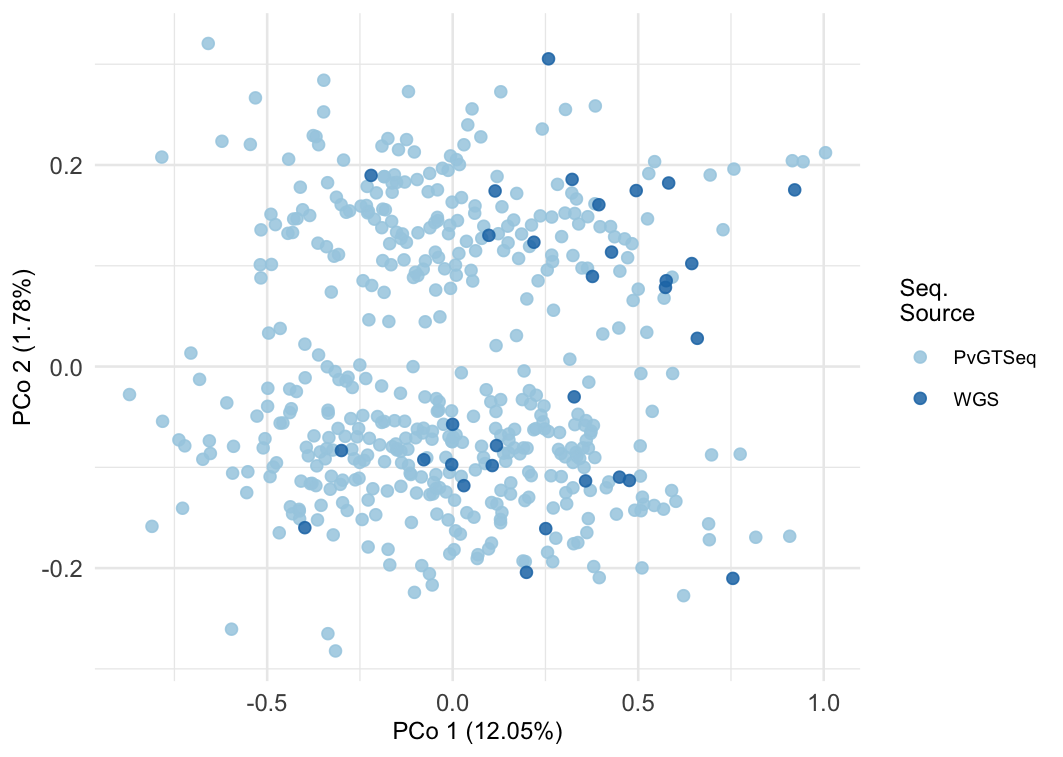

**Fig S13:** Principal coordinate analysis of samples from Guyana generated through PvGTSeq (light blue dots) and WGS (blue dots).
