## Supplementary material for "PvGTSeq and PvCRiSP: two amplicon-based targeted sequencing panels for *Plasmodium vivax*": S3 Text: PvCRiSP protocol: S3_Text.docx

### I. Introduction

This standard operating procedure (SOP) describes the laboratory procedures for high throughput CRiSP library preparation for *Plasmodium vivax* genotyping by 4 highly diverse amplicons: VPS11, RIPR, PIGM, and CG2 related. The 4 amplicons are amplified using high-throughput multiplex PCR and used for downstream genotyping by sequencing on the Illumina platform.

### II. Materials

#### PCR1

- PvCRiSP Primers (S4 Table)
- Nuclease Free Water
- Qiagen Master Mix (206152)
- 1.5 mL Eppendorf Tube
- 96-well semi-skirted PCR plate
- VWR Temporary Seal
- Thermo Clear Adhesive Seal
- Thermo Foil Seal
- p200 Tips
- p20 Tips

#### PCR2

- Unique Dual Indices (UDIs)
- KAPA HiFi 2x Master Mix (7958935001)
- 1.5 mL Eppendorf Tube
- 96-well semi-skirted PCR plate

#### Bead cleanup and library QC

- AmpureXP Beads (A36881)
- PCR Strip Tubes
- Ethanol
- Qubit Reagents (Q33231)
- Qubit Tubes (Q32856)
- Agilent Bioanalyzer Kit (5067-4626)
- Roche qPCR Kit (KK4824)
- Roche Standard 0 (KK4906)
- Roche qPCR Plates (4729692001)
- Roche qPCR Seal (4729757001)

#### Sequencing

- PhiX (FC-110-3001)
- MiSeq v2 Reagent Kit (300 cycles) (MS-102-2002)

### III. Procedure

#### 1. Workflow summary

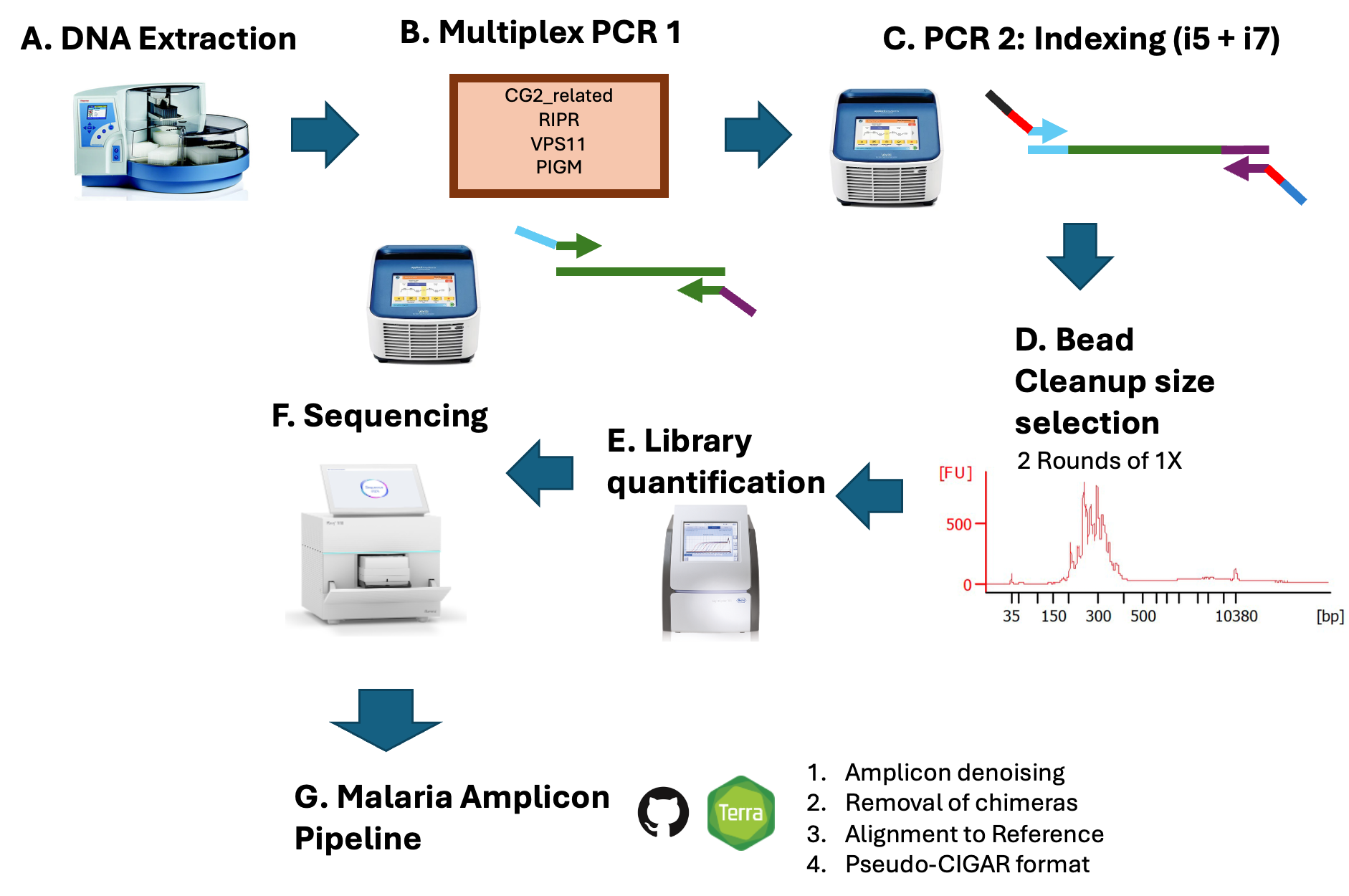

#### 2. PvCRiSP PCR1 (Multiplex PCR)

##### a. Primer re-suspension and preparation of the primer mix and pre-mix

Each primer pair is ordered as a forward and reverse primer that comes lyophilized in individual tubes. Spin down the lyophilized pre-mix and add the required volume of nuclease-free dH_2_O to reach the final concentration of 200 uM for each individual primer. Vortex well, then spin down. Create primer stocks combining the forward and reverse at 100 uM.

##### b. Primer mix preparation

1. To prepare the CRiSP primer stock solution, take an equal amount of each primer pair and combine them together in a 1.5 ml eppendorf tube.
2. Your primer working solution will be at 5 uM. If you want 100 reactions you would add the volumes seen in Table 1 below:

| **Table 1** | |
| --- | --- |
| **Reagent** | **100 Rx Volume (uL)** |
| **100 nM primers (VPS11, RIPR, PIGM, CG2 related)** | 10 |
| **Nuclease-free dH2O** | 120 |

##### c. PvCRiSP PCR1 Master Mix

1. Preparation of master mix (MM):

| **Table 2** | | |
| --- | --- | --- |
| **Reagent** | **Working Solution** | **1Rx Volume (uL)** |
| **Qiagen Plus Master Mix** | 2 X | 10 |
| **Primer Mix** | 5 uM | 2 |
| **Nuclease-free dH2O** | --- | 2 |
| **DNA** | --- | 6 |
| **Total Volume** |  | 20 |

2. Add 14 uL PCR1 MM to each well of the plate.
3. Add 6 uL of sample genomic DNA (or negative, positive control templates). Spin down plate for ~30 seconds.
4. Place plate on a thermocycler with the following amplification settings:

| **Table 3** | | |
| --- | --- | --- |
| **Step** | **Temperature** | **Time** |
| 1 | 95C | 15 Sec |
| 2 | 95C | 30 Sec |
| 3 | 57C | 30 Sec (5% ramp, ~0.3 C/s) |
| 4 | 72C | 2 min |
| 5 | Go to Step 2 | 5 X |
| 6 | 95C | 30 Sec |
| 7 | 65C | 30 Sec |
| 8 | 72C | 30 Sec |
| 9 | Go to Step 6 | 20 X |
| 11 | 4C | Inf |

5. Once the thermocycler completes, dilute PCR1 product 1:10 (10uL product, 90 uL nuclease-free dHO_2_). Mix slowly >20 times with the pipette set to 90uL. Seal the plate and spin down.

#### 3. PvCRiSP PCR2 (Nextera indexing)

1. Prepare PCR2 indexing for each well as follows:

| **Table 3** | |
| --- | --- |
| **Reagent** | **1RX volume** |
| **KAPA HiFi HotStart ReadyMix (2x)** | 10 uL |
| **Unique dual index**** | 4.4 uL |
| **Total** | 14.4 uL |

**Unique dual indices stored in a 10 uM plate; it is best to use distinct index sets when performing sequential seq.

2. Add 6 uL of diluted PCR1 product. Seal and spin down the plate.
3. Place plate on a thermocycler with the following amplification settings:

| **Table 4** | | |
| --- | --- | --- |
| **Step** | **Temperature** | **Time** |
| 1 | 95C | 3 min |
| 2 | 98C | 20 sec |
| 3 | 65C | 30 sec |
| 4 | 72C | 30 sec |
| 5 | Go to Step 2 | 10 X |
| 6 | 72C | 1 min |
| 7 | 4C | Inf |

4. Once PCR2 is complete, combine 15uL of each PCR2 product into a 5 ml Eppendorf tube.

#### 4. Bead cleanup size selection

#### a. AmpureXP bead-based size selection (left-tailed clean-up)

1. Equilibrate AmpureXP beads to room temperature (RT) for 30 minutes prior to use. Similarly, take out the Bioanalyzer reagents out of the 4 °C at this time.
2. Make 5 ml 80% ethanol solution.
3. Aliquot 90 uL of combined, well-mixed PCR2 product into each of 8 tubes within a 0.2 mL tube strip. Each aliquot will receive the same 1X (90uL) AmpureXP bead input volume.
4. Vortex AmpureXP beads thoroughly. Promptly add 90 uL to the 8 sample aliquots. Mix thoroughly using a multichannel pipette set to 180 uL.
5. Incubate at RT for 5 minutes.
6. Place on a magnetic stand until the solution clears (3+ minutes).
7. Carefully discard the supernatant without disturbing the bead pellet.
8. While still on the magnetic stand, add 180 uL of fresh 80% ethanol to the beads and incubate for 30 seconds. Remove the supernatant and discard. Repeat this wash a second time.
9. Leaving the tubes on the magnetic stand, remove any residual ethanol with a small (e.g., p20) pipette and/or allow ethanol to evaporate for 2 minutes. Do not exceed this evaporation time.
10. Remove tubes from the magnetic stand and add 15 uL EB buffer (10 mM Tris-Cl, pH 8.5) directly onto the beads. Mix thoroughly and let incubate at RT for 5 minutes.
11. Place on the magnet once again until the solution clears (3+ minutes).
12. Collect 13 uL of the supernatant without disturbing the pellet into a single 0.2 mL tube and repeat steps 3-11. At the end, collect and combine supernatants into a single, labeled tube for the next part of the protocol.

#### b. BioAnalyzer QC

#### 1. Allow the gel-dye mix to equilibrate to room temperature for 30 minutes before use. Protect the gel-dye mix from light during this time.

#### 2. Take a new High Sensitivity DNA chip out of its sealed bag and place the chip on the chip priming station.

#### 3. Pipette 9.0 μl of the gel-dye mix at the bottom of the well-marked and dispense the gel-dye mix.

#### 4. Set the timer to 60 seconds, make sure that the plunger is positioned at 1 ml and then close the chip priming station. The lock of the latch will click when the Priming Station is closed correctly.

5. Press the plunger of the syringe down until it is held by the clip.
6. Wait for exactly 60 seconds and then release the plunger with the clip release mechanism.
7. Visually inspect that the plunger moves back at least to the 0.3 ml mark.
8. Wait for 5 s, then slowly pull back the plunger to the 1 mL position.
9. Open the chip priming station.
10. Pipette 9.0 μL of the gel- dye mix in each of the wells marked.
11. Pipette 5 μL of green-capped High Sensitivity DNA marker (green) into the well-marked ladder symbol and into each of the 11 sample wells.
12. Pipette 1 μl of the yellow-capped High Sensitivity DNA ladder vial (yellow) in the well-marked with the ladder symbol.
13. In each of the 11 sample wells pipette 1 μL of sample (used wells) or 1 μL of marker (unused wells).
14. Place the chip horizontally in the adapter of the vortex mixer and make sure not to damage the bulge that fixes the chip during vortexing.
15. Vortex for 60 seconds at 2400 rpm.
16. Open the lid of the Agilent 2100 Bioanalyzer.
17. Check that the electrode cartridge is inserted properly and the chip selector is in position.
18. Place the chip carefully into the receptacle. The chip fits only one way.
19. Carefully close the lid. The electrodes in the cartridge fit into the wells of the chip.
20. The 2100 Expert software screen shows that you have inserted a chip and closed the lid by displaying the chip icon at the top left of the Instrument context.
21. Label your samples on the computer chart. Click the Start button in the upper right of the window to start the chip run. The incoming raw signals are displayed in the Instrument context.

#### 22. Library quantification by qPCR

The library eluted from bead clean-up (BC) likely has a concentration of ca. 100 – 2000 nM. We need to bring it down to picomolar range such that it falls within the standard curve used in this qPCR. Therefore, we will create three serial dilutions (1:100, 1:1000, and 1:10000) from the starting BC product with a final volume of 20uL. Then:

1. Dilute the BC to 1:10 and take 2 uL from BC and add 18 uL dH2O. Mix well and repeat this step three more times to create the dilutions 1:100, 1:1000, and 1:10000.
2. Dilute DNA Standard 0 (200 pM) to 20 pM (i.e., take 2 uL and 18 uL dH2O).
3. Prepare qPCR master mix as follows:

| **Table 11** | |
| --- | --- |
| **Reagent** | **1Rx Volume (uL)** |
| **KAPA SYBR FAST qPCR Master Mix (2X)** | 10 |
| **Primer Mix** | 2 |
| **Nuclease-free H2O** | 4 |
| **DNA** | 4 |
| **Total** | 20 |

4. Add 16 uL qPCR master mix to plate columns 1-3 (rows A-H).
5. Add 4 uL of each standard and the library in a plate with three technical replicates.
6. Seal and spin down plate.
7. Place plate on qPCR thermocycler with the following amplification settings:

| **Table 12** | | |
| --- | --- | --- |
| **Step** | **Temperature** | **Time** |
| **Step 1** | 95 C | 5 min |
| **Step 2** | 95 C | 30 secs |
| **Step 3** | 60 C | 45 secs |
| **Step 4** |  | Go to Step 2 35 X |

8. Once the run is finished use the following formulae to calculate the concentration of the library:

$Library [nM]=\frac{Lib. \left[ pM \right]*Lib. Dil. factor*Stock standard 0 \left[ pM \right]*Standard 0 size (bp)}{Standard 0 \left[ pM \right]* Standard 0 dil. factor*Mean Lib.size (bp)}$

#### 23. Sequencing

##### a. Sequencing through iSeq100

###### 1. Thaw the Bagged Cartridge

1. Put on a new pair of powder-free gloves.
2. Remove the cartridge from -25°C to -15°C storage.
3. If the cartridge is boxed, remove it from the box but *do not open the white foil bag.*
4. Thaw the bagged cartridge using one of the following methods. Use immediately after thawing, without refreezing or otherwise storing.

| Table 13 | | |
| --- | --- | --- |
| Method | Thaw Time | Instruction |
| 20°C to 25°C water bath | 6 hours, not exceeding 18 hours | Use 6 L (1.5 gal) water per cartridge. Set a temperature-controlled water bath to 25°C or mix hot and cold water to achieve 20°C to 25°C. Face the bag label up, submerge the cartridge completely, and apply ~2 kg (4.5 lb) weight to prevent floating. Do not stack cartridges in the water bath unless it is temperature-controlled. |
| 2°C to 8°C refrigerator | 36 hours, not exceeding 72 hours | Position the cartridge so that the label faces up and air can circulate on all sides, including the bottom. |
| Room temperature air | 9 hours, not exceeding 18 hours | Position the cartridge so that the label faces up and air can circulate on all sides, including the bottom. |

###### 2. Library Preparation

Prepare the flow cell as follows:

1. Remove a new flow cell from 2°C to 8°C storage.
2. Set aside the unopened package at room temperature for 10–15 minutes.
3. Remove Resuspension Buffer (RSB) from -25°C to -15°C storage. Alternatively, use 10 mM Tris-HCl pH 8.5 or EB in place of RSB.
4. Remove 10 nM PhiX stock from -25°C to -15°C storage.
5. Thaw RSB and PhiX at room temperature for 10 minutes.
6. In a 200 uL low-bind microtube, dilute the library in RSB to the volume of 20 uL and a final concentration of 1nM.

It is possible to store the 1 nM library at -25°C to -15°C for up to 1 month.

7. In a 200 uL low-bind microtube, combine 15 µl of the 1 nM library with 85 µl of RSB to dilute the library to the loading concentration of 150pM.
8. Set aside the diluted library on ice for sequencing. **Sequence libraries the same day they are diluted**.
9. In a 200 uL low-bind microtube, dilute the PhiX in RSB to the volume of 20 uL and a final concentration of 1nM.

It is possible to store the 1 nM PhiX at -25°C to -15°C for up to 1 month.

10. In a 200 uL low-bind microtube, combine 15 µl of the 1 nM PhiX with 85 µl of RSB to dilute the PhiX to the loading concentration of 150pM.
11. Finally combine 95 µl of the Library with 5 µl of the PhiX for a 5% spike-in.

###### 3. Load Consumables Into the iSeq100 Cartridge

1. Open the cartridge bag from the notches.
2. Avoiding the access window on top of the cartridge, remove the cartridge from the bag. Discard the bag.
3. Invert the cartridge five times to mix reagents. Internal components can rattle during inversion, which is normal.
4. Tap the cartridge (label facing up) on the bench or other hard surface five times to ensure reagent aspiration.
5. Using a new pipette tip, pierce the library reservoir and push the foil to the edges to enlarge the hole.
6. Discard the pipette tip to prevent contamination.
7. Add 20 µl diluted library to the *bottom* of the reservoir. Avoid touching the foil.
8. Open the white foil flow cell package from the notches. Use within 24 hours of opening.
9. Pull the flow cell out of the package.
10. Touch only the plastic when handling the flow cell.
11. Avoid touching the electrical interface, CMOS sensor, glass, and gaskets on either side of the glass.
12. Hold the flow cell by the grip points with the label facing up.
13. Insert the flow cell into the slot on the front of the cartridge.

##### b. Sequencing through MiSeq V2

###### 1. Thaw the Bagged Cartridge

1. Thaw the reagent cartridge using a room temperature water bath.

Alternatively, thaw reagents overnight in 2°C to 8°C storage. Reagents are stable up to one week when stored at this temperature.

2. Remove the cartridge from -25°C to -15°C storage.
3. Place the reagent cartridge in a water bath containing enough room temperature deionized water to submerge the base of the reagent cartridge. Do not allow the water to exceed the maximum water line printed on the reagent cartridge.
4. Allow the reagent cartridge to thaw in the room temperature water bath until it is thawed completely.

MiSeq v2 cartridges— ~ 60 minutes.

5. Remove the cartridge from the water bath and gently tap it on the bench to dislodge water from the base of the cartridge. Dry the base of the cartridge.
6. Invert the reagent cartridge ten times to mix the thawed reagents, and then inspect that all positions are thawed.
7. Inspect the reagents in positions 1, 2, and 4 to make sure that they are fully mixed and free of precipitates.
8. Gently tap the cartridge on the bench to reduce air bubbles in the reagents.

The MiSeq sipper tubes go to the bottom of each reservoir to aspirate the reagents, so it is important that the reservoirs are free of air bubbles.

9. Place the reagent cartridge on ice for up to six hours, or set aside at 2°C to 8°C until ready to set up the run. For best results, proceed directly to loading the sample and setting up the run.

###### 2. Library Preparation

1. Combine 800 uL of molecular grade H2O and 200 uL 1N NaOH.

Use the fresh dilution within 12 hours.

2. Remove HT1 from -25°C to -15°C storage and thaw at room temperature. Store at 2C to 8C until you are ready to dilute the library.
3. Remove 10 nM PhiX stock from -25°C to -15°C storage and thaw at room temperature for 10 minutes.
4. In a 200 uL low-bind microtube, dilute the library in 10 mM Tris-HCl pH 8.5 or EB to the volume of 20 uL and a final concentration of 6 nM.

It is possible to store the 6 nM library at -25°C to -15°C for up to 1 month.

5. Denature the 6 nM library. In a 1.5 mL tube combine 5 uL of 6 nM Library with 0.5 uL 0.2N NaOH, vortex and then centrifuge at 280xg for 1 min.
6. Incubate at room temperature for 5 min.
7. Add 990 uL prechilled HT1 Buffer to the tube containing the denatured library. The result is 1 mL of a 30 pM denatured library.
8. Combine 420 uL of 30 pM denatured library with 180 uL HT1 Buffer. The final result is a diluted library to the final concentration of 21 pM and 500 uL volume.
9. Set aside the diluted library on ice for sequencing. **Sequence libraries the same day they are diluted**.
10. In a 200 uL low-bind microtube, dilute the PhiX in 10 mM Tris-HCl pH 8.5 or EB to the volume of 20 uL and a final concentration of 4 nM.
11. Denature the 4 nM PhiX. In a 1.5 mL tube combine 5 uL of 4 nM PhiX with 0.5 uL 0.2N NaOH, vortex and then centrifuge at 280xg for 1 min.
12. Incubate at room temperature for 5 min.
13. Add 990 uL prechilled HT1 Buffer to the tube containing the denatured PhiX. The result is 1 mL of a 20 pM denatured library.
14. Combine 375 uL of 20 pM denatured PhiX with 225 uL HT1 Buffer. The final result is a diluted PhiX to the final concentration of 12.5 pM and 500 uL volume.
15. Finally combine 555 µl of the Library with 45 µl of the PhiX for a 7.5% spike-in.

###### 3. Load consumables into the cartridge

1. Using a new 1 mL pipette tip, pierce the library reservoir and push the foil to the edges to enlarge the hole.
2. Discard the pipette tip to prevent contamination.
3. Add 600 µl diluted library to the reservoir. Avoid touching the foil.
4. Follow instrument instructions to place the cartridge in the machine.
