## Supplementary material for "PvGTSeq and PvCRiSP: two amplicon-based targeted sequencing panels for *Plasmodium vivax*": S2 Text: PvGTSeq protocol: S2_Text.docx

### I. Introduction

This standard operating procedure (SOP) describes the laboratory procedures for high throughput PvGTSeq library preparation for *Plasmodium vivax* genotyping by 249 amplicons. The 249 amplicons are amplified using high-throughput multiplex PCR and used for downstream genotyping by sequencing on the Illumina platform. The 249 amplicons include a panel of 213 amplicons distributed every 200 Kb across the *P. vivax* genome for population differentiation at three geographic scales: Global regions, countries in the Americas, and within countries in the Americas. Additionally, it includes 36 amplicons against 10 antimalarial resistance associated genes.

### II. Materials

#### Selective whole genome amplification (sWGA):

- Primers pvset1
- Primers pvset1920
- GenomiPhi Enzyme/Buffer (25660031)
- Nuclease Free Water
- dNTPs (4mM)

#### PCR1

- PvGTSeq Primers (S4 Table)
- Nuclease Free Water
- Qiagen Master Mix (206152)
- 1.5 mL Eppendorf Tube
- 96-well semi-skirted PCR plate
- VWR Temporary Seal
- Thermo Clear Adhesive Seal
- Thermo Foil Seal
- p200 Tips
- p20 Tips

#### PCR2

- Unique Dual Indices (UDIs)
- KAPA HiFi 2x Master Mix (7958935001)
- 1.5 mL Eppendorf Tube
- 96-well semi-skirted PCR plate

#### Bead cleanup and library QC

- AmpureXP Beads (A36881)
- PCR Strip Tubes
- Ethanol
- Qubit Reagents (Q33231)
- Qubit Tubes (Q32856)
- Agilent Bioanalyzer Kit (5067-4626)
- Roche qPCR Kit (KK4824)
- Roche Standard 0 (KK4906)
- Roche qPCR Plates (4729692001)
- Roche qPCR Seal (4729757001)

#### Sequencing

- PhiX (FC-110-3001)
- MiSeq v2 Reagent Kit (300 cycles) (MS-102-2002)

### III. Procedure

#### A. Workflow summary

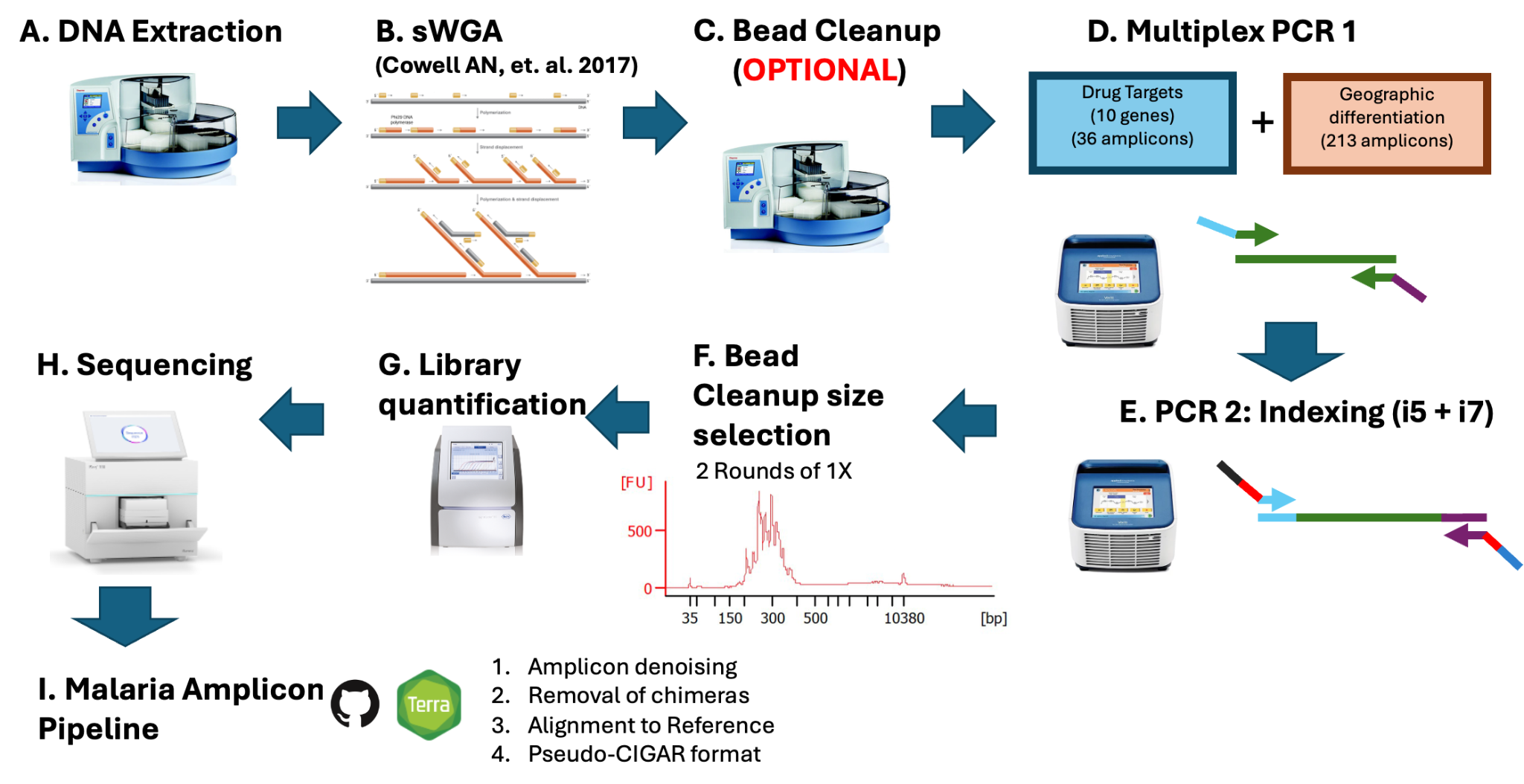

#### B. sWGA of Plasmodium vivax with GenomiPhi v2

##### b. First amplification round

1. Prepare the samples mix following specification of the **Table 1**

| **Table 1** | | | |
| --- | --- | --- | --- |
| **Reagent** | **Working Solution** | **Final Concentration** | **1Rx Volume (uL)** |
| **pvset1920 [uM]** | 250 | 6.5 | 0.52 |
| **dNTPs [mM]** | 4 | 1.25 | 6.25 |
| **H2O** |  |  | 0.23 |
| **DNA** |  |  | 3 |

2. Heat the samples to 95C for 3 minutes then cool to 4C on ice.

Heating the DNA for longer than 3 minutes or at higher temperatures can cause damage to the DNA.

3. Prepare Reaction mix following specifications on table 2:

| **Table 2** | |
| --- | --- |
| **Reagent** | **1Rx Volume (uL)** |
| **Reaction buffer (Blue)** | 9 |
| **Enzyme Phi29 (Yellow)** | 1 |

Prepare the master mix only in sufficient quantities and **immediately prior to use**. Keep the master mix on ice and discard any unused portion. The master mix contains all the components required for DNA amplification and will generate **off-target amplification products if exposed to temperatures > 4°C for sufficient time.**

4. Transfer the Reaction mix to the cooled Sample mix.
5. Incubate for DNA amplification following instructions on **Table 3**:

| **Table 3** | | |
| --- | --- | --- |
| **Step** | **Temperature** | **Time** |
| **1** | 35C | 5 min |
| **2** | 34C | 10 min |
| **3** | 33C | 15 min |
| **4** | 32C | 20 min |
| **5** | 31C | 30 min |
| **6** | 30C | 60 min |
| **7** | Got to step 6 | 23 X |
| **8** | 65C | 10 min |
| **9** | 4C | Hold |

##### c. Second amplification round

1. Dilute the first-round product to 1/50 with molecular grade H2O.
2. Prepare Reaction mix following specifications on **Table 4**:

| **Table 4** | | | |
| --- | --- | --- | --- |
| **Reagent** | **Working Solution** | **Final Concentration** | **1Rx Volume (uL)** |
| pvset1 [uM] | 250 | 3.4 | 0.272 |
| dNTPs [mM] | 4 | 1.25 | 6.25 |
| H2O |  |  | 0.478 |
| 1st round DNA product |  |  | 3 |

3. Heat the samples to 95C for 3 minutes then cool to 4C on ice.
4. Transfer the Reaction mix to a cooled Sample mix.
5. Incubate for DNA amplification following instructions on **Table 3.**
6. Dilute the final product in 1/50 with molecular grade H_2_O (**RECOMMENDED**) or Bead clean up with the protocol AmpureXP sWGA bead clean-up with MagPin.

#### C. AmpureXP sWGA bead clean-up with MagPin (OPTIONAL)

1. Equilibrate the AmpureXP beads to room temperature for 30 minutes and make a fresh 80% ethanol solution.
2. Prepare the wash plates and the elution plate (volumes above).
3. Transfer 20 uL of sWGA product to a new plate.
4. Add 36 uL of beads (1.8 x ratio) – pipette to mix.
5. Incubate at room temperature for 5 minutes.
6. Make sure there is a clean Cover Plate on the magnet - place into the sample plate so that the beads bind.
7. Incubate for 3 minutes – it is sometimes helpful to gently swirl the magnet to make sure that most of the beads are picked up.
8. Move the magnet (w/beads attached) to the first wash plate containing 100 uL of 80% ethanol.
9. Allow the magnet to sit for 30 seconds.
10. Move the magnet to the second wash plate containing 100 uL of 80% ethanol.
11. Allow the magnet to sit for 30 seconds.
12. Transfer the magnet w/beads to a new plate - dry for 1 minute.
13. Move the magnet to the elution plate containing 30 uL of low TE buffer.
14. Remove the Cover Plate from the magnet – swirl the Cover Plate make sure the beads come off the plastic and into solution.
15. Pipette to mix, incubate for 5 minutes.
16. Add the magnet back - incubate for 3 minutes.
17. Remove the magnet and discard the Cover Plate/beads.
18. Leaves 30 uL of supernatant containing the cleaned product.
19. Repeat steps 16 and 17 if a lot of beads remain.

#### D. PvGTSeq PCR1 (Multiplex PCR)

##### a. Primer Re-suspention and preparation of the primer mix and pre-mix

Each pair of primer comes in a pooled version in a 96 plate format, where in each well it is a pair of primers. Spin down the lyophilized pre-mix, and add the required volume of nuclease-free dH_2_O to reach the final concentration of 200 uM for each individual primer (See table 5). It’s very important to mix thoroughly (pipette set to high volume) and vortex briefly to ensure the pellet is completely solubilized. Then, spin down.

| **Table 5** | |
| --- | --- |
| Pool Concentration | 40 nmol |
| Number of primers in pool | 2 |
| Individual Primer concentration | 20 nmol |
| Final concentration of each primer | 200 uM |
| Resuspension volume | 100 uL |

In the table above the initial concentration of each individual primer is 20 nmol (40 mol in total in the well), so the re-suspension volume for each well is 100 uL to reach a final concentration of 200 uM of each individual primer.

##### b. Primer pre-mix preparation

1. The primers are grouped and sorted in 4 sets based on their final concentration in the PCR reaction. Final concentration of each primer is described in S3 Table.
2. To prepare each pre-mix set, take an equal amount of each primer pair in the set and combine them together in a single micro-tube. The minimum required **volume of each primer pair** in the primer set pre-mix for 100 reactions is in the 6th column of the table 5.

| **Table 6** | | | | | |
| --- | --- | --- | --- | --- | --- |
| **Primer set** | **Number of primers in the set** | **Primer mix working solution [uM]** | **Final PCR concentration [nM]** | **Required stock of indv. Pairs volume** | **Required pre-mix volume** |
| Set 1 | 16 | 0.067 | 10 | 0.1 | 1.6 uL |
| Set 2 | 61 | 0.1 | 15 | 0.15 | 9.15 uL |
| Set 3 | 119 | 0.2 | 30 | 0.3 | 35.7 uL |
| Set 4 | 53 | 0.3 | 45 | 0.45 | 23.85 uL |
| Total number of primers | **249** |  |  |  |  |
| H_2_O Volume | | | |  | 229.7 uL |

Based on the table above, for 100 reactions (Table PCR Master Mix) it is required to combine 0.1 uL of the stock each primer pair in the first set to prepare 1.6 uL of Set 1 pre-mix; 0.15 uL of each primer pair for the Set 2; 0.3 uL for the set 3, and 0.45 for the set 4. These volumes represent the minimum required volumes, and it is possible to use more if the volumes are too small to pipette, but it is important to use the same amount of each primer pair in the preparation of each set.

##### c. Primer Mix

1. Once all pre-mixes are prepared, combine them in a single tube following the instructions in the 5th column on the table **Primer Sets**. Thus the required volume for 100 reactions for the Sets 1, 2, 3 and 4 are 1.6 uL, 9.15 uL, 35.7 uL and 23.85 uL respectively.
2. Finally, add 229.7 uL of dHO_2_ to have the primers at the working concentration of 0.0667, 0.1, 0.2, 0.3 uM for primers in the primer sets 1, 2, 3, and 4 respectively.

All these numbers are to prepare 300 uL of primer mix that is required for 100 reactions

##### d. PvGTSeq PCR1 Master Mix

1. Preparation of master mix:

| **Table 7** | | |
| --- | --- | --- |
| **Reagent** | **Working Solution** | **1Rx Volume (uL)** |
| **Qiagen Plus Master Mix** | 2 X | 10 |
| **Primer Mix** | 0.067 - 0.3 uM | 3 |
| **Nuclease-free dH2O** | --- | 1 |
| **DNA** | --- | 6 |
| **Total Volume** |  | 20 |

2. Add 14 uL PCR1 cocktail to every plate well.
3. Add 6 uL of sample genomic DNA (or negative/positive control templates). Spin down plate for ~30 secs.
4. Place plate on a thermocycler with the following amplification settings:

| **Table 8** | | |
| --- | --- | --- |
| **Step** | **Temperature** | **Time** |
| 1 | 95C | 15 Sec |
| 2 | 95C | 30 Sec |
| 3 | 57C | 30 Sec (5% ramp, ~0.3 C/s) |
| 4 | 72C | 2 min |
| 5 | Go to Step 2 | 5 X |
| 6 | 95C | 30 Sec |
| 7 | 65C | 30 Sec |
| 8 | 72C | 30 Sec |
| 9 | Go to Step 6 | 20 X |
| 11 | 4C | Inf |

5. In a clean plate, aliquot 130 uL of nuclease-free dHO_2_.
6. Create 1/13 PCR1 product dilution by adding 120 uL of the aliquoted nuclease-free dHO_2_ directly to PCR1 product (10 uL). Mix **slowly** >20 times (pipette set to high volume). Seal the plate and spin down.

#### E. PvGTSeq PCR2 (Nextera indexing)

1. Prepare PCR2 indexing as follows:

| **Table 9** | |
| --- | --- |
| **Reagent** | **1RX volume** |
| **KAPA HiFi HotStart ReadyMix (2x)** | 5 uL |
| **2.2 uL unique dual index**** | 2.2 uL |
| **Total** | 7.2 uL |

**Unique dual indices stored in a 10 uM plate; it is best to use distinct index sets when performing sequential seq.

2. Add 3 uL of diluted PCR1 product. Seal and spin down the plate.
3. Place plate on a thermocycler with the following amplification settings:

| **Table 10** | | |
| --- | --- | --- |
| **Step** | **Temperature** | **Time** |
| 1 | 95C | 3 min |
| 2 | 98C | 20 sec |
| 3 | 65C | 30 sec |
| 4 | 72C | 30 sec |
| 5 | Go to Step 2 | 10 X |
| 6 | 72C | 1 min |
| 7 | 4C | Inf |

For MiSeq, pool 2 plates of 96 samples each for a total of 192 samples (ensure that different index plates were used for the two plates). For iSeq pool 64 samples (including positive and negative controls) for each sequencing run.

4. Combine 7 uL of each PCR2 product into a 1.5 ml Eppendorf tube. Discard the remaining PCR product. Avoid combining the whole PCR product as it can generate bias in the read depth per sample because of heterogeneous evaporation on the plate.

#### F. Bead clean-up size selection

##### a. *AmpureXP* bead-based size selection (left-tailed clean-up)

1. Equilibrate AmpureXP beads to room temperature for 30 min prior to using them. Additionally, take BioAnalyzer reagents out of 4 °C at this time.
2. Make 5 ml 80% ethanol solution.
3. Aliquot 50 uL of combined, well-mixed PCR2 product into each of 8 tubes within a 0.2 mL tube strip. Each of these 8 aliquots will receive a similar 1X (50uL) AmpureXP bead input volume.

**Note:** The volume of 50 uL is for combining 64 samples. If a full plate is run, increase the starting volume of the aliquots and the AmpureXP beads to 90 uL.

4. Vortex AmpureXP beads thoroughly. Promptly add 50 uL to the 8 sample aliquots. Mix thoroughly.
5. Incubate at room temperature for **5 min**.
6. Place on a magnetic stand until the solution clears **(3+ min)**.
7. Discard the supernatant without disturbing the bead pellet.
8. While still on the magnetic stand, add 180 uL fresh 80% ethanol to the beads and incubate for 30 seconds.  Remove supernatant and discard. Repeat this wash once.
9. While still on the magnetic stand, remove any residual ethanol with a small (e.g., p20) pipette and/or allow ethanol to evaporate for **2 min** (with tubes uncovered). **Do not exceed 2 min evaporation time.**
10. Remove from the magnetic stand and add 15 uL EB buffer (10 mM Tris-Cl, pH 8.5). Mix thoroughly and let incubate at room temperature for **5 min**.
11. Place on the magnetic stand until the solution clears **(3+ min)**.
12. Collect 13 uL supernatant without disturbing the pellet.
13. Combine 10 uL of all supernatants in a single 0.2 mL tube and repeat the selection using 80 uL of AmpureXP beads.

#### I. Malaria amplicon pipeline

Paired-end Illumina sequencing data is processed in the form of FASTQ files using a custom analysis pipeline for which documentation can be found at:

<https://github.com/broadinstitute/malaria-amplicon-pipeline>.

This pipeline utilizes the Divisive Amplicon Denoising Algorithm (DADA2) to obtain microhaplotypes (i.e., alleles or ‘amplicon sequence variants’). Then, microhaplotypes obtained from DADA2 are aligned against a custom-built database of PvP01 reference sequences for each amplicon locus. We then summarized observed sequence polymorphism into a concise format by converting individual microhaplotypes into ‘pseudo-CIGAR’ strings using a custom python script that can be found at:

<https://github.com/Paulonvnv/MHap-Analysis/blob/main/docs/functions_and_libraries/ASV_to_CIGAR2.py>

The rules used for this conversion are also detailed in Fig S2 in S1 Text.

Next, microhaplotypes were discarded if supported by fewer than 10 read-pairs or by less than 1% of total read-pairs within a heterozygous locus. We also filtered out microhaplotypes exhibiting less than 80% identity to the reference sequence. We then masked variants in both the FASTA sequence of the microhaplotype and its pseudo-cigar string representation according to three criteria: 1) SNPs or INDELs located in homopolymer regions (≥5 consecutive identical nucleotides), 2) INDELs positioned at either the beginning or end of the microhaplotype sequence, and 3) variants predominantly (> 66%) present as minor alleles and mainly found in heterozygous sites (> 66%) across the population. A detailed description of these filters are described in the following rMarkdown document:

<https://github.com/Paulonvnv/PvGTSeq_PvCRiSP_paper/blob/main/RMD_Reports/Sequencing_performance_and_filtering.Rmd>

All downstream analyses were performed based on the pseudo-cigar strings using custom R scripts that can be found at <https://github.com/Paulonvnv/PvGTSeq_PvCRiSP_paper>, and those scripts are coordinated using the following rMarkdown document:

<https://github.com/Paulonvnv/PvGTSeq_PvCRiSP_paper/blob/main/Draft_PvGTSeq_paper.Rmd>

All functionalities created to handle pseudo-cigar strings are documented at <https://github.com/Paulonvnv/MHap-Analysis>. This pipeline is also implemented in a Terra workspace —a user-friendly cloud-based platform for processing amplicon sequencing data, and the documentation of this workspace can be found at:

<https://publichealth.terra.bio/#workspaces/malaria-featured-workspaces/Malaria_Plasmodium_Illumina_Amplicon>

And its GitHub repository can be accessed using the following link:

<https://github.com/broadinstitute/malaria>
